# Can measurements of inflammatory biomarkers be used to spot viral infections?

**DOI:** 10.1101/2020.10.06.20207860

**Authors:** Anirban Sinha, René Lutter, Tamara Dekker, Barbara Dierdorp, Peter J. Sterk, Urs Frey, Edgar Delgado-Eckert

## Abstract

Accurate detection of human respiratory viral infections is highly topical. We investigated how strongly inflammatory biomarkers (FeNO, eosinophils, neutrophils, and cytokines in nasal lavage fluid) and lung function parameters change upon rhinovirus-16 infection, in order to explore their potential use for infection detection. To this end, within a longitudinal cohort study, healthy and mildly asthmatic volunteers were experimentally inoculated with rhinovirus-16 and time series of these parameters/biomarkers were systematically recorded and compared between the pre- and post-infection phases of the study, which lasted 2 and 1 month/s, respectively. We found that the parameters’/biomarkers’ ability to discriminate between the infected and the uninfected state varied over the observation time period. Consistently over time, the concentration of cytokines in nasal lavage fluid showed moderate to very good discrimination performance, thereby qualifying for disease progression monitoring. On the other hand, lung function and FeNO, while quickly and non-invasively measurable using cheap portable devices (e.g., at airports), performed poorly.

## 1. Introduction

Infections of the respiratory tract in humans are a major cause for global morbidity and mortality. About 80% of such infections are caused by viruses, the most prevalent being influenza, parainfluenza, respiratory syncytial virus (RSV), coronavirus, adenovirus, and rhinovirus [1]. Early detection and accurate identification of the virus in potentially affected individuals are necessary for appropriate and timely remedies and management. However, the differential diagnosis can be difficult due to the non-specific clinical presentation and symptoms [2], particularly in premorbid patients with, e.g., bronchial asthma. Historically, examination of clinical symptoms in combination with virus isolation, serological tests, imaging, and chest radiography [3], have served as mainstays of viral detection [4]. Later, chest computed tomography [5], antibody staining and detection methods complemented with immune assays paved the way for better diagnostics but still lacked the specificity and sensitivity required for accurate detection.

In the last two decades, there has been a remarkable improvement in the diagnosis of respiratory pathogens with the availability of molecular assays. Indeed, mainly due to the advent of polymerase chain reaction (PCR) techniques, highly sensitive and robust methods such as nucleic acid sequence-based amplification (NASBA), loop-mediated isothermal amplification (LAMP), and microRNAs [1,6-8] have come into the picture. Multiplex assays coupled to microfluidics and high-resolution imaging constitute further developments towards more accurate diagnostics [1]. These latest diagnostic tools rely on either finding the virus or traces of it in the patient’s body, or virus specific antibodies generated by the patient’s immune system. A topical example is SARS-CoV-2. Currently, the most accurate diagnosis method to detect SARS-CoV-2 infections in humans is based on detecting amplified nucleic acid sequences of the virus genome [9]. However, PCR technology may not be fully accurate [10,11] and often requires multiple testing for reliable results [12].

On the other hand, clinically available parameters for airways assessment such as inflammatory and lung function indices have not been traditionally used for the diagnosis of viral respiratory infections. This is mainly due to the fact that pathological changes in such indices have been found to lack pathogen-specificity [13,14]. Furthermore, to the best of our knowledge, the discrimination accuracy of such parameters for the detection of any viral respiratory infection in humans, regardless of the specific virus involved, has never been systematically tested. Whether a physiological parameter or biomarker qualifies for the detection of a viral respiratory infection depends on how accurately it can discriminate between infected and uninfected states. This can only be the case if the change in the parameter/biomarker elicited by the infection is stronger than its natural fluctuations during uninfected phases. Moreover, ideally, this should be independent of other underlying disease processes, such as chronic ailments.

Therefore, we aimed at investigating how reliable inflammatory biomarkers and lung function are for the detection of respiratory viral infections in humans, by systematically comparing the values of these parameters/biomarkers during the participants’ pre- and post-infection state within a longitudinal study cohort. Our approach consisted of using longitudinally measured inflammatory biomarkers and lung function parameters to assess whether post-infection values exceeded their natural variability during uninfected phases. Because of ethical reasons, we focused on experimental infection of humans in vivo with rhinovirus 16 (RV16), a not particularly virulent pathogen causing the common cold.

## 2. Materials and Methods

### 2.1. Study cohort and parameters/biomarkers measured

In a prospective, longitudinally designed study comprising 12 healthy and 12 mildly asthmatic subjects, we measured time series of a set of inflammatory/immune biomarkers and standard lung function [15]. The cohort demographics have been previously published in [15] and are also summarized again in Table 1 below. The serum antibody titer of RV16 required had to be less than 1:8 for all participants on the day of enrollment in order to exclude any participants whose immune system had already been exposed to the virus. Based on our previous findings, a serum antibody titer of less than 1:6 is sufficient to exclude such participants [16]. However, to increase recruitment of participants and without compromising the validity of the study outcomes, a relaxed titer of 1:8 was chosen. The participants were within 18-30 years of age and without concomitant disease or pregnancy. Asthmatic subjects were not on steroids at least in the previous 6 weeks before study screening.

**Table 1.** The cohort demographics. BMI is Body Mass Index. Only 1 healthy subject smoked 2 pack years or less 2 years before recruitment to our study, which is considered an insignificant smoking history. * mean ± standard deviation, PY: Pack-year.

| Demographic features | Healthy | Asthmatic |
| --- | --- | --- |
| Total number, n | 12 | 12 |
| Female gender, n (%) | 7 (58.3%) | 8 (66.7%) |
| Age (years), mean (SD) | 21 ± 1.5 | 22.2 ± 2.2 |
| Ethnicity (Caucasian), n (non-Caucasian, n) | 11 | 9 |
| BMI, mean (SD)* | 22.2 ± 1.6 | 22.8 ± 3.1 |
| Smoking (pack years), n | 1 (0.17 PY) | -- |
| Height (centimeters)* | 177.7 ± 8.6 | 172.5 ± 13.0 |
| Weight (KG)* | 70.4 ± 10.1 | 67.8 ± 12.4 |
| Baseline spirometry measurements |  |  |
| FEV1 %predicted* | 105.7 ± 11.6 | 101.0 ± 10.0 |
| FVC %predicted* | 104.2 ± 10.5 | 104.2 ± 10.2 |
| PEF %predicted* | 108.4 ± 14.0 | 104.7 ± 12.2 |

The inclusion/exclusion criteria for participants along with the study design have been published in detail before [15]. A summary of inclusion criteria can be found in Table 2 below.

**Table 2.** Study inclusion characteristics for volunteers. AHR: Airway Hyper Responsiveness, PC20: Provocative Concentration causing a 20% fall in FEV1), SPT: Skin Prick Test.

| <u>Healthy</u> | <u>Asthmatics</u> |
| --- | --- |
| No history of episodic chest symptoms | History of episodic chest symptoms |
| Baseline FEV1 ≥ 80 % predicted | Baseline FEV1 ≥ 70% predicted |
| AHR to methacholine (PC<sub>20</sub>) ≥ 19.6 mg/ml | AHR to methacholine (PC<sub>20</sub>) ≤ 9.8 mg/ml |
| SPT negative for all 12 common Aeroallergens | SPT positive for at least 1 out of 12 common Aeroallergens |

The parameters screened were cellular immune-markers including eosinophil and neutrophil cell numbers in nasal lavage fluid. Molecular immune markers, also measured in nasal lavage fluid, were 10 cytokines: IFN-gamma, IL-1β, IL-10, IL-13, IL-17A, IL-33, IL-6, IL-8, IP-10, and TNF-alpha, which are biomarkers known to play a role in asthma, inflammation, and RV16 infection [17]. The inflammatory biomarker, measured in the exhaled air, was nitric oxide (FeNO). The lung function parameters measured were Peak Expiratory Flow (PEF), Forced Expiratory Volume in 1st second (FEV1), Forced Vital Capacity (FVC), and the ratio FEV1/FVC.

These parameters/biomarkers were repeatedly captured during two months prior to and one month following an experimental RV16 infection induced by controlled and deliberate inoculation with RV16 of healthy and mildly asthmatic volunteers [15]. The sampling frequencies of the various parameters/biomarkers are detailed below in Table 3. In order to minimize the influence of diurnal variations of the cytokines measured in nasal lavage fluid, for any given participant, the sampling visits were scheduled in such a way that samples were collected at approximately the same time of the day.

**Table 3.** Overview of the different samples collected from study participants and their frequency of sampling before and after rhinovirus challenge.

| **Biomarker or parameter** | **Sampling frequency before rhinovirus challenge** | **Sampling frequency after rhinovirus challenge** |
| --- | --- | --- |
| Lung function (FEV1, FVC, FEV1/FVC, PEF) | 2x daily | 2x daily |
| Exhaled Nitric Oxide (FeNO) | 3x weekly | 3x weekly |
| Eosinophil and neutrophil cell numbers in nasal lavage fluid | 1x weekly | 3x weekly |
| Cytokines in nasal lavage fluid | 1x weekly | 3x weekly |

The study protocol was approved by the hospital medical ethical committee. All study participants provided written informed consent of participation. The cohort study has been registered in the Netherlands Trial Register, NL5317 (NTR5426).

The efficacy of the inoculation with RV16 was established using serum antibody tests (RV16 seroconversion), clinical symptoms, and rhinovirus Polymerase Chain Reaction (PCR) conducted on nasal lavage fluid taken from every participant after the inoculation. The viral load just after a few days from rhinovirus challenge along with the seroconversion for antibodies against rhinovirus (measured at the end of the study for each participant) are summarized in Table 4 below. See also Supplement for symptoms data. In addition, the development of a host response was carefully assessed for each cohort participant as described previously. All these tests provided strong evidence supporting the fact that all 24 participants were effectively infected with the RV16 after inoculation [15]. All participants were continuously monitored throughout the study. Clinical symptoms and occurrence of any concomitant infection due to any exposures to pathogens apart from the experimental challenge were carefully recorded using a detailed questionnaire (see Supplement), which was filled out at every visit. Indeed, during the pre-challenge phase, none of the participants reported symptoms of any concomitant infection that could possibly confound the results during this phase of the study. Apart from that, PCR tests using primers from a panel of common respiratory viruses were performed for every individual before experimental challenge to rule out the occurrence of concomitant infections.

**Table 4:** Overview of the viral load [copies/mL] measured in nasal lavage fluid from each participant. Samples of nasal lavage fluid were collected on days 1, 3 and 6 after challenge. NA means no sample was collected. The rightmost column displays, for each participant, the results of the antibody test against RV (from seroconversion) performed at the last visit of the study (approximately 30 days after challenge). 0 = No response, 1= Response.

| Subject ID | Viral load in nasal lavage fluid measured using RVPCR (copies/mL) |  |  | Seroconversion response |
| --- | --- | --- | --- | --- |
|  | Day 1 after challenge | Day 3 after challenge | Day 6 after challenge |  |
| 01A | 4040 | 95600 | 8520 | 1 |
| 02A | 138000 | 0 | 4970 | 0 |
| 04A | 0 | 0 | 0 | 1 |
| 05A | NA | 359000 | 486000 | 0 |
| 06A | 18000 | 1350000 | 14500 | 1 |
| 07A | NA | 1580000 | 216000 | 1 |
| 08A | 0 | 0 | 0 | 1 |
| 09A | NA | 62700 | 1240000 | 0 |
| 10A | 0 | NA | 0 | 1 |
| 11A | 3210 | 4310000 | 3690000 | 1 |
| 12A | NA | 86900 | NA | 0 |
| 13A | NA | 0 | NA | 0 |
| 01H | NA | 29300 | NA | 0 |
| 03H | NA | 1180000 | 13000 | 0 |
| 05H | 18300 | 9640000 | 13300 | 1 |
| 06H | 0 | 32700 | 0 | 0 |
| 07H | 0 | 0 | 0 | 1 |
| 08H | 0 | 26900 | NA | 1 |
| 09H | 0 | 169000 | 7690000 | 1 |
| 11H | 0 | 0 | 0 | 0 |
| 12H | 2560 | 15700 | 7160 | 1 |
| 13H | 0 | 239000 | 2630000 | 1 |
| 14H | 0 | 16600000 | 13600000 | 0 |
| 15H | 0 | 0 | 220000 | 1 |

### 2.2. Computational methods and statistical analysis

For every inflammatory/immune/lung functional biomarker separately, we calculated the average value over the first 10 days after inoculation and compared it to the average calculated over every possible time interval of 10 consecutive days prior to the inoculation. Previously published work suggests that all important pathophysiological and immunological changes elicited by a rhinovirus infection in humans are most likely to happen within a time interval of 10 days after exposure. This constitutes the rationale behind our choice of time interval or “window” length [18].

In order to assess the time dependency of the above-mentioned comparison of averages, the 10-day-windows prior to the inoculation were glided, one day at a time. The start position of the gliding window is expressed relative to the day of the viral inoculation, which is marked as day 0. Chronologically speaking, the first pre-inoculation window considered in our analysis starts on day - 50 relative to the day of the inoculation, and the last one at position −9 (the last window contains the day of the inoculation). See Figure 1 below for more details.

**Figure 1.**
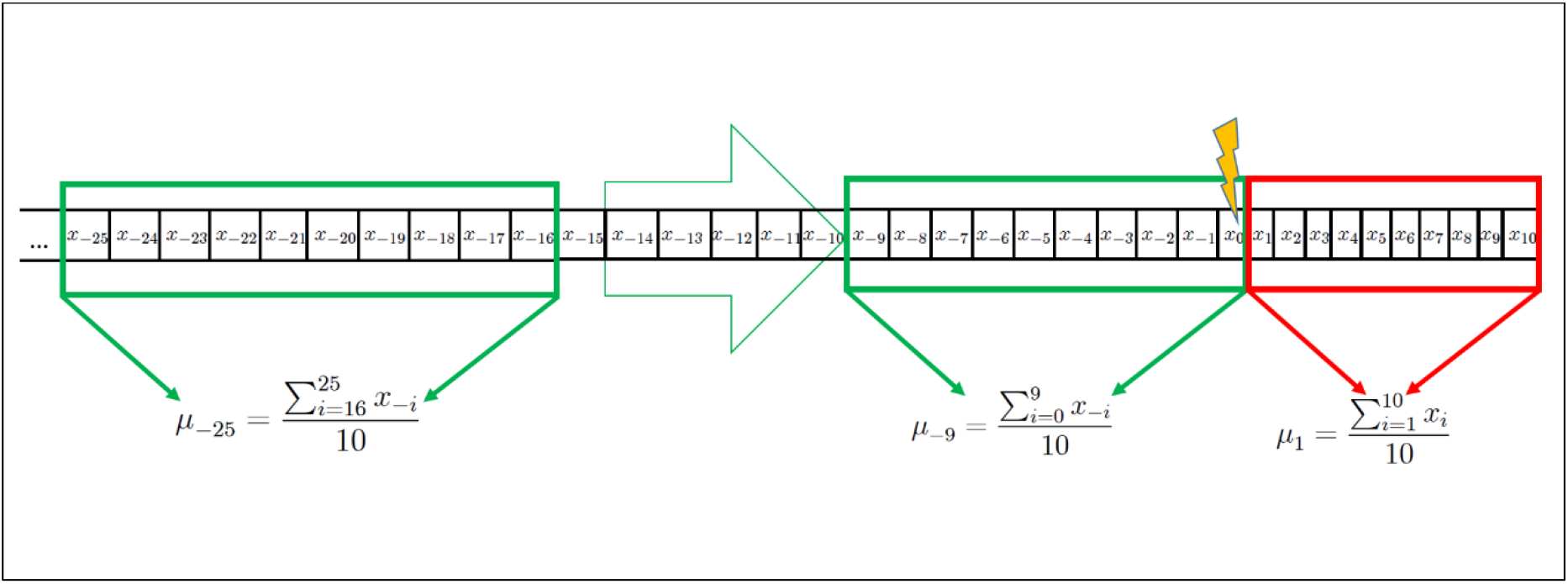
Graphical representation of a participant’s biomarker time series x_i_. The chronological order of measurements of the biomarker is expressed in days from the day of inoculation, which is marked as “day 0”. Negative indexes represent measurements taken before the viral challenge, whereas positive indexes belong to measurements conducted after the challenge. Note that for those biomarkers not measured on a daily basis, some of the values in the time series are missing. A moving window (depicted in green), covering ten days, glides, one day at a time, during the pre-challenge phase, starting from day −50 down to day −9. For each window, the average over those ten days the window covers is calculated. For example, the window starting on day −25 yields the average μ_-25_. After the challenge, only one window is contemplated, namely the window covering the first ten days immediately after inoculation. This window yields the average μ_1_. For every participant, there is such an average μ_j_ and μ_1_, respectively, where *j* = −50,…,-9. For a given day *j* between −50 and −9 during the pre-challenge phase, and for a given subgroup of the cohort (healthy, asthmatics, and pooled), the empirical distribution of averages μ_j_ were compared to the empirical distribution of averages μ_1_ using a two-tailed paired Mann-Whitney-U-test. The results (p-values) of these comparisons are depicted for the biomarkers IL-6 and FeNO in the top panels of Figures 2 and 3 below. Summary statistics for the pooled group can be found in Tables “Summary statistics of the averages of IL-6 and FeNO concentration within each window” in Supplementary data. Furthermore, ROC curves were constructed assuming that the collection of averages μ_j_ is characteristic of uninfected individuals, and the collection of averages μ_1_ is characteristic of infected individuals. The areas under these ROC curves (AUC) are depicted for the biomarkers IL-6 and FeNO in the bottom panels of Figures 2 and 3 below.

The above-mentioned comparison of within-window-averages was not done individually, but rather for all cohort participants using a two-tailed paired Mann-Whitney-U-test. Thus, the test aims at establishing whether the distributions of values of a given parameter/biomarker prior to and during the infection are significantly different. For a given pre-inoculation window, the p-value resulting from this test is indicative of the ability of a given inflammatory/immune or lung function biomarker to discriminate between healthy and infected states as a function of the starting position of the pre-inoculation window at hand (see Top Panel in Figures 2 and 3). Thereby, we looked at how the range observed during the healthy state varies in time and whether it remains, over time, statistically distinct from the range observed during the infected state. Furthermore, this discriminatory ability was also quantitatively assessed by constructing a receiver operating characteristic (ROC) curve [19] and calculating the area under the same (see Bottom Panel in Figures 2 and 3). The area under a ROC curve (AUC) is an effective way to summarize the overall discrimination accuracy of a given parameter/biomarker at a given time point during the pre-challenge phase. If there is no apparent distributional difference between the values of the parameter/biomarker for a given pre-challenge window and the values for the post-challenge window, the AUC will have a value of about 0.5. On the other hand, if there is a perfect separation of the values for a given pre-challenge window and the values for the post-challenge window, the AUC will be equal to 1. To relate the AUC to its discriminatory performance, we follow [20], and use the following denominations of AUC ranges: 0.9– 1.0 excellent; 0.8-0.9 very good; 0.7-0.8 good; 0.6-0.7 sufficient; 0.4-0.6 bad. This is how we quantitatively compared the discrimination accuracy of a given parameter/biomarker at a given time point during the pre-challenge phase to its discrimination accuracy at another time point during the pre-challenge phase (Bottom Panel in Figures 2 and 3). Moreover, we used the AUC to compare two different parameters/biomarkers in terms of their discrimination accuracy. For this comparison, however, it was also important to consider the stability over time of the discrimination accuracy of each of the two biomarkers.

**Figure 2.**
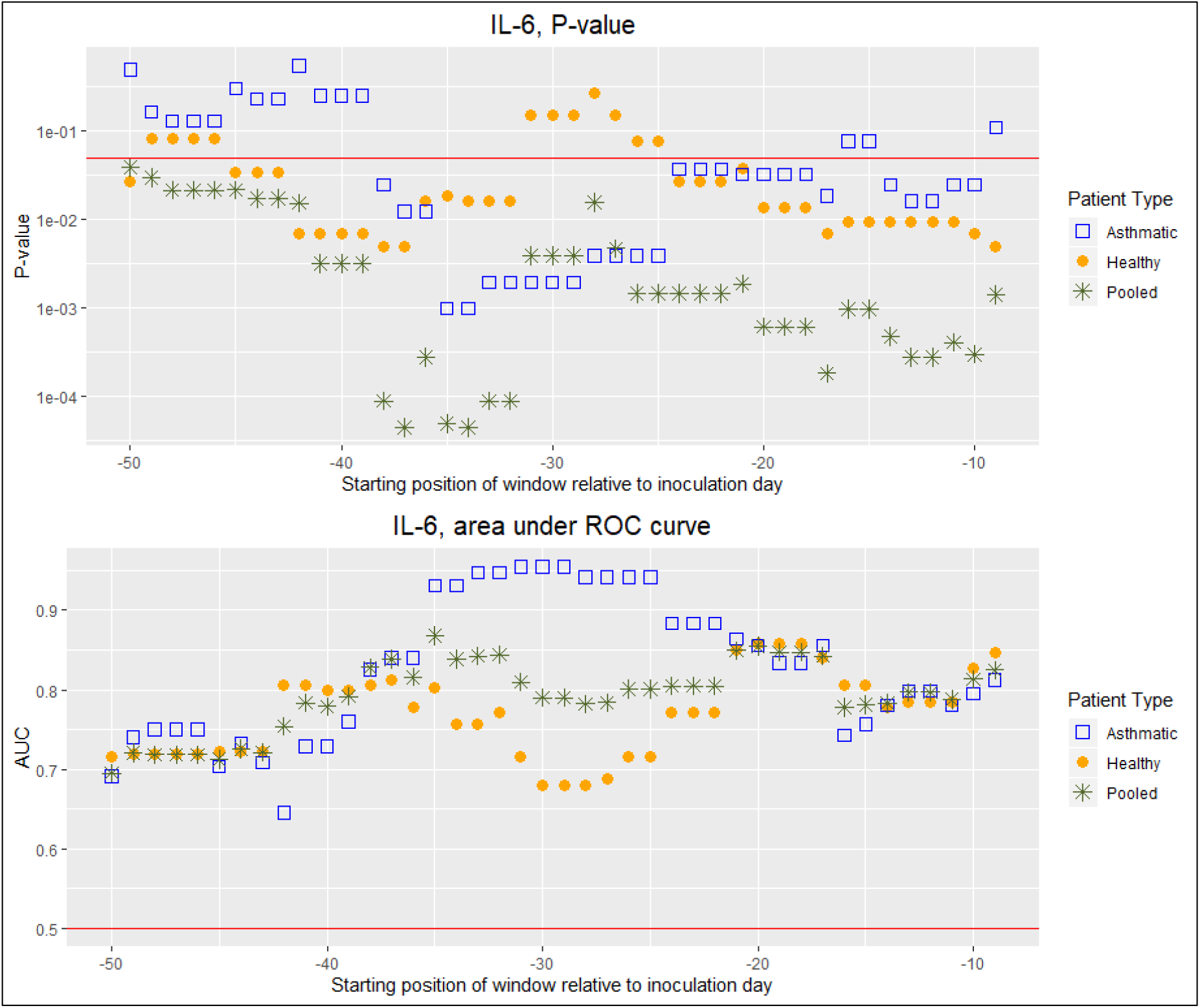
Top Panel: P-value resulting from two-tailed paired Mann-Whitney-U-test as a function of the starting day of the gliding 10-days-window expressed relative to the day of viral inoculation (“day 0”). Each test compares the average concentration of IL-6 in nasal lavage fluid calculated over the first 10 days after inoculation to the average calculated over the time interval of 10 consecutive days starting on the day prior to the inoculation indicated on the horizontal axis. The red horizontal line indicates the significance level of 0.05. Vertical axis uses a logarithmic scale. Bottom Panel: Area under the curve (AUC) as a function of the starting day of the gliding 10-days-window expressed relative to the day of viral inoculation (“day 0”). Each AUC value corresponds to the ROC curve that assesses the discrimination power of the average concentration of IL-6 in nasal lavage fluid calculated over the first 10 days after inoculation when compared to the average calculated over the time interval of 10 consecutive days starting on the day prior to the inoculation indicated on the horizontal axis. The red horizontal line indicates the average AUC of a completely random classifier (0.5).

**Figure 3.**
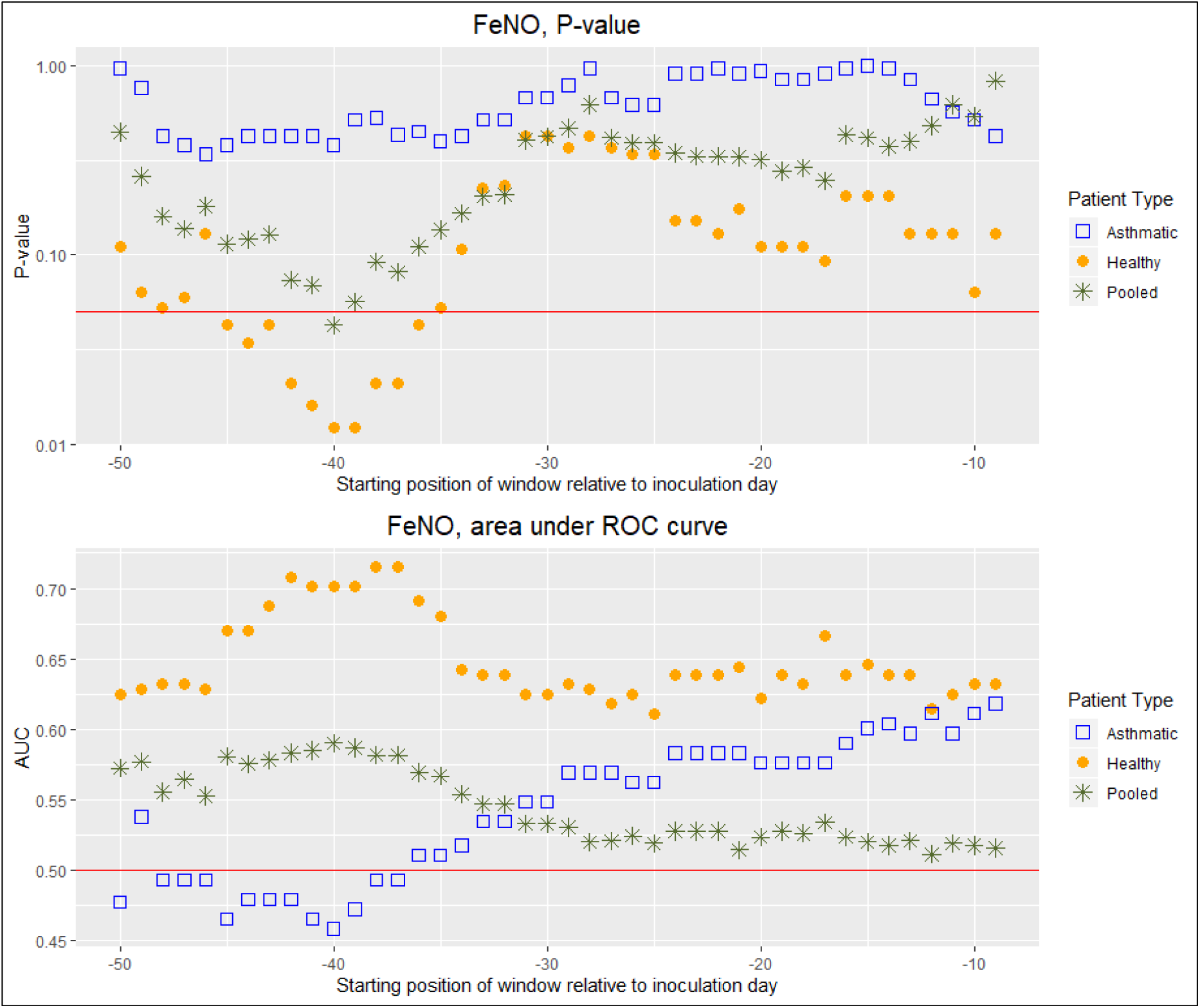
Top Panel: P-value resulting from two-tailed paired Mann-Whitney-U-test as a function of the starting day of the gliding 10-days-window expressed relative to the day of viral inoculation (“day 0”). Each test compares the average FeNO calculated over the first 10 days after inoculation to the average calculated over the time interval of 10 consecutive days starting on the day prior to the inoculation indicated on the horizontal axis. The red horizontal line indicates the significance level of 0.05. Vertical axis uses a logarithmic scale. Bottom Panel: Area under the curve (AUC) as a function of the starting day of the gliding 10-days-window expressed relative to the day of viral inoculation (“day 0”). Each AUC value corresponds to the ROC curve that assesses the discrimination power of the average FeNO calculated over the first 10 days after inoculation when compared to the average calculated over the time interval of 10 consecutive days starting on the day prior to the inoculation indicated on the horizontal axis. The red horizontal line indicates the average AUC of a completely random classifier (0.5).

In order to explore whether the discrimination performance of the parameters/biomarkers at hand is independent of other underlying disease processes, such as chronic ailments, the above described analysis was conducted on the groups of asthmatics and healthy individuals separately. However, we also carried out the analysis on the data resulting from pooling these two groups.

## 3. Results

The cytokines IL-6 (Fig.1, see also **“**Summary statistics of the averages of IL-6 concentration within each window” in Supplementary data for summary statistics of the pooled group), IFN-gamma, IL-10, IL-13, IL-8, and TNF-alpha (Figures S1(A,B)-S5(A,B) in SD) measured in nasal lavage fluid display AUC values fluctuating between 0.6 and 0.95. This makes these biomarkers at least sufficient, if not good, very good, or excellent discriminators throughout the observation period. IP-10, IL-1β, IL-17A, and IL-33 show a similar pattern with an overall lower discrimination performance, including several time intervals of bad performance (AUC values 0.55 to 0.9, with IL-13 occasionally falling into the interval 0.4-0.5; see Figures S6(A,B)-S9(A,B) in SD).

For cellular immune-markers measured in nasal lavage fluid the emerging picture is more complex (see SD). The eosinophil cell numbers were found to be a sufficient discriminator (AUC values 0.6 to 0.7), although bad during limited time intervals. Whereas for neutrophil cell numbers this appeared to be the other way around, with some episodes of good discrimination performance (Figures S10(A, B) and S11(A, B) in SD). Surprisingly, the overall cell numbers in nasal lavage fluid oscillates between being a sufficient and a good to very good discriminator (Figure S12(A, B) in SD).

FeNO is, in general, a bad discriminator (AUC 0.5-0.6), except for non-asthmatic patients, for which it is sufficient, and good only during a limited time interval (Fig.2, see also **“**Summary statistics of the averages of FeNO concentration within each window” in Supplementary data for summary statistics of the pooled group).

The lung function parameters are throughout bad discriminators, whether measured in the morning or in the evening (See Figures S13(A, B)-S20(A,B) in SD for the occasional exceptions to this rule).

A general pattern observed in our results is how the ability to discriminate between healthy and RV16-infected states varied over time. The amplitude of these temporal fluctuations was found to be relatively highest among the cellular immune-markers and some of the cytokines.

When comparing asthmatics and non-asthmatics in terms of the ability of the various parameters/biomarkers to discriminate between healthy and RV16-infected, it appeared that for FeNO and the lung function parameters, with the exception of PEF and the ratio FEV1/FVC, the discriminatory performance was higher in the non-asthmatics group. This difference between the two groups could be observed over time intervals of weeks (See Figure 3 and Figures S13(A, B)-S20(A,B) in SD). For the cellular immune-markers and the cytokines, however, discriminatory performance differences were less pronounced or less consistent over time (See Figure 2 and Figures S1(A,B)-S12(A,B) in SD).

## 4. Discussion

### 4.1 Main findings and interpretations

To the best of our knowledge, this is the first study in which the utility of inflammatory/immune biomarkers and lung function has been tested for detecting respiratory viral infections, while considering the natural temporal fluctuations of the parameters/biomarkers studied.

Our findings indicate that the concentrations of the various cytokines and the overall cell count in nasal lavage fluid are consistently at least sufficient discriminators of RV infection, the cytokines at times even excellent discriminators. Furthermore, eosinophil and neutrophil cell numbers in nasal lavage fluid were found to reach, at times, the level of a sufficient discriminator. However, this was not consistently the case throughout the observation period. Finally, FeNO and the lung function parameters were, in general, bad discriminators, except within the group of non-asthmatics, where sufficient discrimination power was reached during certain time intervals of the observation period.

Within a period of time of 7 weeks prior to experimental and controlled inoculation with RV-16, each cohort participant was closely monitored and no symptoms of any respiratory disease were reported during this time interval. Nevertheless, our analysis has revealed a time dependency of the discriminative power of all the parameters/biomarkers studied herein. This can be interpreted in two ways: 1) During the phase prior to inoculation, the parameters/biomarkers fluctuate with an amplitude large enough to occasionally reach or nearly reach values characteristic of an infection. 2) The effect elicited by the RV-16 infection on these parameters/biomarkers is not strong enough, and thus the values remain within a “normal” range that can be observed during infection-free phases. In the context of discriminating between the non-infected and the infected state, the first interpretation hints at the possibility of finding false positives, whereas the second interpretation indicates that false negatives may result.

Whilst FeNO and lung function parameters can be quickly and non-invasively measured using cheap portable devices (e.g., at airports), our results suggest that these parameters do not qualify as accurate discriminators of RV infection. On the other hand, we found that the concentrations of the various cytokines in nasal lavage fluid may indeed be successfully used for the detection of RV infection. In particular, IL-6 (Fig.1) and IFN-gamma (Figure S1(A,B) in SD) are the best discriminating cytokines, despite the natural temporal fluctuations in their concentrations. This may not be surprising, as these cytokines are known to play critical roles in local anti-viral responses [21]. Furthermore, the influence of the asthma condition on IFN-gamma’s discrimination performance is almost negligible.

What is the clinical implication of these data? A nasal lavage is a relatively bothering procedure that needs to be carried out by trained health care workers. Moreover, the laboratory assays to measure the concentration of these cytokines are expensive and require proficient staff and laboratory facilities. Consequently, we conclude that measuring these cytokines as biomarkers in nasal lavage fluid will be most suitable in a hospital setting. In fact, some cytokines have been shown to be good predictors of disease progression in flu patients [22].

### 4.2 Limitations of this study

Our data analysis strategy of averaging biomarkers over a sliding 10-days window has the, for our purposes undesirable effect of smoothing the time series data, thereby artificially reducing the true variability of the parameters/biomarkers. In fact, moving average filters are a widely used time series smoothing technique [23]. However, on which days after rhinovirus infection a given patient will display the strongest changes in the parameters/biomarkers measured in our study is highly variable from patient to patient. Previous research work indicates that the strongest changes will typically occur within the first 10 days after infection [18]. Therefore, we used the average of a given parameter/biomarker over the first ten days after inoculation as the magnitude that would be representative of the changes occurring upon infection, without having to choose a particular day after infection that may not be suitable for all individuals. Consequently, and for the sake of consistency, we also used averages of the parameters/biomarkers over 10-day windows prior to inoculation.

In previous work [15] we explored the potential of the whole parameter/biomarker time series to characterize each participant’s state upon inoculation. Those findings suggest that a “personalized time-series analysis” would most likely have more power and lead to better discriminatory performance. However, such an approach is challenging from a practical point of view, as patients would need to be constantly monitored, let’s say weekly, in order to detect a potential infection no one knows in advance when it will hit. This would constitute a big burden for most patients or healthy individuals, which discouraged us from pursuing such an approach in the context of rhinovirus infection detection. Nevertheless, it is conceivable that, in the future, there will be portable patient tele-monitoring technologies that allow for passive patient monitoring, i.e., longitudinal measuring of relevant parameters/biomarkers without the patient’s active participation. With such technologies a personalized time-series analysis approach will indeed become feasible. However, we believe that the development of such technologies is still in early stages. Furthermore, and more importantly, we think that the potential ethical issues arising from constantly tele-monitoring patients or healthy individuals need to be traded-off against the presumptive health benefits of such technologies.

The choice of the age group for our study volunteers (18-35 years) was determined by the fact that in this study the participants were inoculated with an active virus. Indeed, ethical approval by the hospital board for the inclusion of pediatric or older patients would have been very difficult to obtain. Our results may have been influenced by the presence of atopy/allergy in our cohort. However, our study was designed to mimic a real-life setting in which the majority of the asthma patients have atopy as opposed to non-atopic asthma [24]. Moreover, some of the cytokines we decided to measure in this study (e.g., IFN-g) are known for their increased secretion during rhinovirus infections and less for their appearance during allergic reactions. Furthermore, we selected mediators that would be released at sufficient abundancy in the nasal compartment to enable detection, even in the absence of any respiratory infection, and that have been consistently reported in the field [25,26].

Although our study has an important clinical and epidemiological message, rhinovirus infections represent a milder form of respiratory infection in comparison to other more virulent pathogens. Hence, the conclusions derived from this study can only serve as potential reference for other viruses infecting the human respiratory system. Moreover, the sample size of our study is relatively small owing to the number of sampling visits every volunteer was subjected to. However, this shortcoming was compensated by the unprecedented exceptionally high sampling frequency of parameters/biomarkers.

## Supporting information

Copy of Study Questionnaire

Supplemental Figures S1 through S20

## Data Availability

The following are available online on medRxiv: Figures S1 through S20 displaying the same information as Figures 2 and 3 in the manuscript, but concerning all the remaining parameters/biomarkers used in this study. Tables T1 through T22 displaying summary statistics of the averages, within each gliding time window, of each parameter/biomarker used in this study. Furthermore, Table T23 displaying symptoms data upon inoculation for each study participant, and a copy of the study questionnaire filled out for each participant at each visit. For every parameter/biomarker, and for each cohort participant, pre-challenge time series of windowed averages can be made available upon email request to the corresponding author.

## Supplementary Materials

The following are available online: Figures S1 through S20 displaying the same information as Figures 2 and 3 above, but concerning all the remaining parameters/biomarkers used in this study. Tables T1 through T22 displaying summary statistics of the averages, within each gliding time window, of each parameter/biomarker used in this study. Furthermore, Table T23 displaying symptoms data upon inoculation for each study participant, and a copy of the study questionnaire filled out for each participant at each visit. For every parameter/biomarker, each cohort participant’s pre-challenge time series of windowed averages can be used by researchers as reference values to compare to values measured on their own patients infected with similar/different viruses known to affect the respiratory system. These time series can be made available upon request.

## Author Contributions

Conceptualization: AS, UF, RL, PJS, TD and BD; methodology: AS, EDE, UF, RL and PJS; formal analysis: EDE and AS; data curation: EDE and AS; coding: EDE; manuscript writing: EDE and AS; supervision: UF, RL and PJS.; project administration: UF, RL and PJS, EDE and AS; funding acquisition: UF, RL and PJS and AS. All authors have read and agreed to the published version of the manuscript.

## Acknowledgments

We would like to acknowledge all the participants who volunteered to participate in our study along with all master students, research technicians and study nurses who helped us in parts of the project.

