## Supplementary material for "Can measurements of inflammatory biomarkers be used to spot viral infections?": Copy of Study Questionnaire

### Worksheet for BIOFLUC

(Version date: 03 NOV 2015)

**Protocol number: NL54293.018.15**
**METC number: 2015_226**
**NTR(TC= 5426)**

Temporal fluctuations of biomarkers in patients with asthma and controls:
Proof of concept for predicting loss of control

*Confidential*

Subject Screening N°: .. .. .. .. .

Investigators: prof.dr. P.J. Sterk , dr. R. Lutter
Primary Investigator: prof.dr. P.J. Sterk

Academic Medical Center
University Hospital of Amsterdam
Department of Pulmonology
Meibergdreef 9, Room F5-147
NL-1105 AZ Amsterdam, the Netherlands

#### Instructions for using this worksheet

- This version is designed for use in BIOFLUC (study protocol NL54293.018.15)

- This worksheet consists of the following sections:
  Visit 1: Screening
  Visit 2: Run in (Day -14)
  Visit 2: Baseline (Day 0)
  Visit 4-23: Follow-up (Day 2 – Day 47)
  Visit 24: RV16-Challenge (Day 49)
  Visit 25-35: Post RV16- Challenge Follow-up (Day 51 – Day 74)
  End of Study
  Data entry check form

- Go through each page of this worksheet. Record Subject Screening N° and/or Subject Allocation N° on each page of the worksheet and on any companion forms (e.g. In-/Exclusion form, Adverse Events form, Medication form)

- Upon completion of the study the Investigator will sign the Data entry check form, after verifying that the filled-in data are complete and accurate.

- All procedures are to be performed according to instructions provided in the protocol or in the latest version of the Procedure manual. Any deviation from the procedure methods described in these documents MUST be documented and explained on this worksheet or appropriately documented elsewhere.

- Choose from available answers by marking the empty box with a tick or cross (√ or X) or by deleting what is not applicable.

- The operators conducting the procedure must indicate their initials next to each completed procedure

- Record the Visit Date on the first page of each visit and the Time of data/material collection next to the procedure on the worksheet (where applicable). If one or more procedures are performed on another day, or an update is required, record the correct Date and Time separately in the Initials box next to the applicable procedure.

- Use the following format for the Date:
  -Two digits for the Day
  - The first three characters of the Month
  - Four digits for the Year

- Use the 24 h format for the Time

- A change or correction of already filled-in data has to be dated and signed. If data has to be corrected, blot it out (e.g. ~~Correction~~). Under all circumstances the original data are not to be masked.

**VISIT 1: SCREENING**

| ITEM | Initials | Data / Comments |
|---|---|---|
| Visit date: | | _ _ / _ _ _ / _ _ _ _ |
| **INFORMED CONSENT**<br>*signed/dated prior to study procedures* | | <u>Steps taken as part of informed consent process</u>:<br><br>☐ NL54293.018.15 study information (ICF,dd.06NOV2015) was *emailed / provided (indicate which is applicable)* to the subject on: _ _ / _ _ _ / _ _ _ _<br>☐ NL54293.018.15 study information was discussed thoroughly with subject<br>☐ Subject was allowed time to ask questions and all questions were answered<br>☐ The ICF was read and reviewed by the subject<br>☐ Subject signed ICF<br>☐ Subject was given copies of signed ICF<br>☐ Subject was given Subject Identification Card<br>☐ No procedures were performed before the subject signed consent |
| **IN-/EXCLUSION CRITERIA**<br>*Follow protocol section 4.2/4.3* | | Check items that are known at visit 1 on the In-/Exclusion criteria form |
| **DEMOGRAPHICS** | | Date of birth: _ _ /_ _ _ /_ _ _ _<br>(Date of birth indicated in InForm: _ _ /_ _ _ /_ _ _ _)<br><br>Gender: Male / Female<br>If female: ☐ childbearing potential<br>☐ non-childbearing potential (date of last menses: _ _ /_ _ _ /_ _ _ _)<br><br>Ethnicity: ☐ Hispanic or Latino<br>☐ Not Hispanic or Latino<br>Race: ☐ White<br>☐ Black<br>☐ Asian, specify: …………………………….<br>☐ Other, specify: ……………………………. |
| **VOLUNTEER** | | Subject is: ☐ Healthy volunteer<br><br>☐ Mild-moderate asthmatic volunteer using only rescue medication (SABA) – no ICS<br>Date of most recent occurrence: _ _ /_ _ _ /_ _ _ _ |

| | | |
|---|---|---|
| **SMOKING**<br>*status/history including Pack years* | | History of smoking? Yes / No<br>If yes, stopped smoking since: _ _ /_ _ _ /_ _ _ _<br>............................................ x ................................<br>Pack years = No. cigarettes smoked/day x No. of years smoked / 20 = ............. |
| **ALCOHOL** | | Does the subject consume alcohol? Yes / No<br>If yes, specify the number of alcoholic drinks per day: ………………. |
| **CAFFEINE**<br>*coffee, tea, cola, energy drinks, caffeinated beverage* | | Does the subject consume caffeinated beverages? Yes / No<br>If yes, specify the number of caffeinated beverages per day: ………. |
| **MEDICAL HISTORY** | | Were any significant medical history reported? Yes / No<br>If yes, check body system, describe abnormality, indicate start and stop date (or whether the abnormality is ongoing at study entry)<br>☐ Eyes, ears, nose & throat ……………………………<br>☐ Respiratory ……………………………<br>☐ Cardiovascular ……………………………<br>☐ Gastrointestinal ……………………………<br>☐ Genitourinary ……………………………<br>☐ Musculoskeletal ……………………………<br>☐ Neurologic ……………………………<br>☐ Endocrine & metabolic ……………………………<br>☐ Haematopoietic/lymphatic ……………………………<br>☐ Dermatologic ……………………………<br>☐ Psychiatric ……………………………<br>☐ Allergic/immunologic ……………………………<br>☐ Reproductive/breast …………………………… |
| **MEDICATION/ THERAPY HISTORY**<br>*All medication including asthma medication at entry (screening).* | | Does the subject use medication? Yes / No<br>If yes, record prior medication taken by the subject within 6 months before starting the trial on Medication form. |

| | | |
|---|---|---|
| **PHYSICAL EXAMINATION** | | Status of body system? **N**ormal / **A**bnormal / **N**ot **D**one<br><br>Check body system and, if **A**bnormal, describe abnormality below; if **N**ot **D**one, describe why<br><br>**N / A / ND**<br>Skin □ / □ / □ ……………………………………………………<br>Eyes □ / □ / □ ……………………………………………………<br>Ears, nose, throat □ / □ / □ ……………………………………………………<br>Head, neck, thyroid □ / □ / □ ……………………………………………………<br>Heart □ / □ / □ ……………………………………………………<br>Lung □ / □ / □ ……………………………………………………<br>Chest (inc breasts) □ / □ / □ ……………………………………………………<br>Abdomen □ / □ / □ ……………………………………………………<br>Lymph nodes □ / □ / □ ……………………………………………………<br>Musculoskeletal □ / □ / □ ……………………………………………………<br>Neurological □ / □ / □ …………………………………………………… |
| **VITAL SIGNS**<br>*After at least 5 min in a semi-recumbent position* | . . : . . | Height: _ _ _ cm Weight: _ _ _ . _ kg BMI: _ _ , _ kg/m²<br><br>Temperature: _ _ , _°C (tympanic) Respiratory rate: _ _ breaths/min<br><br>Pulse: _ _ _ beats/min Blood pressure (Sys/Dias): _ _ _ / _ _ _ mmHg |
| **SKIN PRICK TEST**<br>*(see prints out for results)* | . . : . . | Result? Positive/Negative |

| | | |
|---|---|---|
| **SPIROMETRY**<br>*Perform pre-bronchodilator measurements* | . . : . . | Did the subject <u>use</u> any short-acting bronchodilators (eg albuterol/salbutamol or ipratropium bromide) within 4 h of testing? Yes / No<br><br>If Yes:<br>1/ specify type and dose of medication used:<br>………………………………………………………………………………<br>2/ specify when the last dose of medication was administered:<br>Date: _ _/_ _ _/_ _ _ _<br>Time: . . : . .<br><br>☐ Spirometry print out available<br><br>Best FEV1: _ , _ _ L %pred: _ _ _ %<br><br>Best FVC : _ , _ _ L % pred: _ _ _ %<br><br>FEV1/FVC x 100% = _ _ _ % |
| **METHACOLINE CHALLENGE** | . . : . . | Result? Positive/Negative<br><br>PC20:_ _ _ |
| **BLOODSAMPLING for Safety tests and RV16/IgE Ab**<br>*(see print outs for lab results)* | . . : . . | ☐ *Blood for Proteomics/Lipidomics/Metabolomics and hRV16 titre* (1x 10 ml Serum)<br>☐ *Blood for Standard Complete Blood Count* (1x 4 ml EDTA) |
| **REVIEW (S)AE** | | (S)AE's? Yes / No<br><br>If yes, record on (S)AE form |
| **REMARKS** | | |

| | | |
|---|---|---|
| **ELIGIBILITY** | | Is the Subject eligible for Baseline visit?<br>☐ Yes<br>☐ No |
| **Register in eCRF only when subject is eligible for baseline visit** | | **TRIAL URL:** https://www.data.castoredc.com/#studies |

**VISIT 2: RUN IN (Day -14)**

| ITEM | Initials | Data / Comments |
|---|---|---|
| Visit date: | | _ _ / _ _ _ / _ _ _ _ |
| **REVIEW (S)AE** | | Change in (S)AE's since last visit? Yes / No<br>If yes, record on (S)AE form |
| **REVIEW CONCOMITANT MEDICATION** | | Change in medication since last visit? Yes / No<br>If yes, record on Medication form |
| **IN-/EXCLUSION CRITERIA**<br>*Follow protocol section 4.3/4.3* | | Complete checklist on In-/Exclusion criteria form |
| **PULMONARY/NASAL EXAMINATION** | | Status of body system? **N**ormal / **A**bnormal<br>Check body system and, if **A**bnormal, describe abnormality below<br>**N / A**<br>Nose □ / □ ……………………………………………<br>Lung □ / □ …………………………………………… |
| **VITAL SIGNS**<br>*After at least 5 min in a semi-recumbent position* | . . : . . | Temperature: _ _ , _°C (tympanic) Respiratory rate: _ _ breaths/min<br>Pulse: _ _ _ beats/min Blood pressure (Sys/Dias): _ _ _ / _ _ _ mmHg |
| **FOT**<br>(see also prints out) | . . : . . | Resistance at 5Hz: _ _ _ _ cmH2O/L*s<br>Reactance at 5Hz: _ _ _ _ cmH2O/L*s<br>Resistance at 11Hz: _ _ _ _ cmH2O/L*s<br>Reactance at 11Hz: _ _ _ _ cmH2O/L*s<br>Resistance at 19Hz: _ _ _ _ cmH2O/L*s<br>Reactance at 19Hz: _ _ _ _ cmH2O/L*s<br>R5 – R19: _ _ _ _ cmH2O/L*s |
| **FeNO** | . . : . . | Has subject refrained from eating, drinking and strenuous exercise for at least 1 hr and consumed green vegetables within 24 hrs? Yes / No<br>_ _ _ ppb |

| | | |
|---|---|---|
| **VOCs_ GS-MS**<br>(see alsoTube Identification Form) | <br>. . : . .<br>. . : . .<br>. . : . . | Has subject refrained from eating, drinking and taking inhalation medication for at least 2 hours? Yes / No<br>Start 5 min tidal breathing<br>Collection of exhaled air in Tedlar bag<br>Adsorption of exhaled air sample on Tenax tubes |
| **VOCs_ SpiroNose**<br>(see also prints out Form) | . . : . . | Did the subject rinse the mouth 3 times with water? Yes / No<br>Has the subject taken alcohol 12 hours before visit to the clinic?<br>Yes/No |
| **Exhaled Breath Condensate**<br>(see prints out for lab results) | . . : . . | Did the subject rinse the mouth before the exam? |
| **NASAL LAVAGE**<br>(see Virology print outs for lab results) | . . : . . | _ _ , _ ml obtained after instillation of 8 ml 0.9% NaCl in R / L nostril |
| → **DISPENSE**<br>(Diary and home airflow monitoring device) | | Explain the use of Diary and compliance<br>Explain the use of home Airflow Monitoring device |
| **URINESAMPLING**<br>*(see print outs for lab results)* | . . : . . | ☐ Urine sample collected |
| **REMARKS** | | |
| **Register in eCRF only when subject is eligible for baseline visit** | | **TRIAL URL:** https://www.data.castoredc.com/#studies |

**VISIT 3: BASELINE (Day 0)**

| ITEM | Initials | Data / Comments |
|---|---|---|
| Visit date: | | _ _ / _ _ _ / _ _ _ _ |
| **REVIEW (S)AE** | | Change in (S)AE's since last visit? Yes / No<br>If yes, record on (S)AE form |
| **REVIEW CONCOMITANT MEDICATION** | | Change in medication since last visit? Yes / No<br>If yes, record on Medication form |
| **IN-/EXCLUSION CRITERIA**<br>*Follow protocol section 4.2/4.3* | | Complete checklist on In-/Exclusion criteria form |
| **PULMONARY/NASAL EXAMINATION** | | Status of body system? **N**ormal / **A**bnormal<br>Check body system and, if **A**bnormal, describe abnormality below<br>**N / A**<br>Nose □ / □ ............................................................<br>Lung □ / □ ............................................................ |
| **VITAL SIGNS**<br>*After at least 5 min in a semi-recumbent position* | . . : . . | Temperature: _ _ , _°C (tympanic) Respiratory rate: _ _ breaths/min<br>Pulse: _ _ _ beats/min Blood pressure (Sys/Dias): _ _ _ / _ _ _ mmHg |
| **FOT**<br>(see also prints out) | . . : . . | Resistance at 5Hz: _ _ _ _ cmH2O/L*s<br>Reactance at 5Hz: _ _ _ _ cmH2O/L*s<br>Resistance at 11Hz: _ _ _ _ cmH2O/L*s<br>Reactance at 11Hz: _ _ _ _ cmH2O/L*s<br>Resistance at 19Hz: _ _ _ _ cmH2O/L*s<br>Reactance at 19Hz: _ _ _ _ cmH2O/L*s<br>R5 – R19: _ _ _ _ cmH2O/L*s |
| **FeNO** | . . : . . | Has subject refrained from eating, drinking and strenuous exercise for at least 1 hr and consumed green vegetables within 24 hrs? Yes / No<br>_ _ _ ppb |

| | | |
|---|---|---|
| **VOCs_ GS-MS**<br>(see alsoTube Identification Form) | <br>. . : . .<br>. . : . .<br>. . : . . | Has subject refrained from eating, drinking and taking inhalation medication for at least 2 hours? Yes / No<br>Start 5 min tidal breathing<br>Collection of exhaled air in Tedlar bag<br>Adsorption of exhaled air sample on Tenax tubes |
| **VOCs_ SpiroNose**<br>(see also prints out Form) | . . : . . | Did the subject rinse the mouth 3 times with water? Yes / No<br>Has the subject taken alcohol 12 hours before visit to the clinic?<br>Yes/No |
| **Exhaled Breath Condensate**<br>(see prints out for lab results) | . . : . . | Did the subject rinse the mouth before the exam? |
| **NASAL LAVAGE**<br>*(see Virology print outs for lab results)* | . . : . . | _ _ , _ ml obtained after instillation of 8 ml 0.9% NaCl in R / L nostril |
| **NASAL SWABS**<br>*(always check NOT to select the nostril used for the nasal lavage)* | . . : . . | ☐ Collected in R / L nostril<br>☐ Not required |

| | | |
|---|---|---|
| **SPUTUM INDUCTION**<br>*Perform FEV1 measurements post salbutamol and after each nebulisation period* | . . : . . | Post-salbutamol FEV1 (L): _ , _ _ (90% FEV1(L): _ , _ _ )<br>(80% FEV1(L): _ , _ _ ) |
| | . . : . . | $O_2$ Saturation (pre-induction): _ _ _ % |
| | | Post-bronchodilator FEV1 **<**50%predicted / **≥**50% predicted *****<br>Post-bronchodilator FEV1 **<**1.0 L / 1.0 – 1.5 L / **≥** 1.5 L ***** |
| | . . : . . | Start try-out 2 min nebulisation period with 0.9% NaCl |
| | | FEV1 (L): _ , _ _ Fall ≤10% / 10-20% / >20%? * Sputum? Yes / No |
| | . . : . . | Start 1st 5 min nebulisation period with 0.9% / 3% / 4.5% NaCl |
| | | FEV1 (L): _ , _ _ Fall ≤10% / 10-20% / >20%? * Sputum? Yes / No |
| | . . : . . | Start 2nd 5 min nebulisation period with 0.9% / 3% / 4.5% NaCl |
| | | FEV1 (L): _ , _ _ Fall ≤10% / 10-20% / >20%? * Sputum? Yes / No |
| | . . : . . | Start 3rd 5 min nebulisation period with 0.9% / 3% / 4.5% NaCl |
| | | FEV1 (L): _ , _ _ Fall ≤10% / 10-20% / >20%? * Sputum? Yes / No |
| | . . : . . | Start 4th 5 min nebulisation period with 0.9% / 3% / 4.5% NaCl |
| | | FEV1 (L): _ , _ _ Fall ≤10% / 10-20% / >20%? * Sputum? Yes / No |
| | | Total time of induction: _ _ min |
| | . . : . . | $O_2$ Saturation (post-induction): _ _ _ % |
| | | * select saline concentration to be used based on (changes in) FEV1 |
| **URINESAMPLING**<br>*(see print outs for lab results)* | . . : . . | ☐ Urine sample collected |
| **BLOODSAMPLING**<br>*(see print out for CBC lab result)* | . . : . . | Has subject fasted for at least 6 hours? Yes / No<br>☐ *Blood for Proteomics/Lipidomics/Metabolomics and hRV16 titre* (1x 10 ml Serum)<br>☐ *Blood for Standard Complete Blood Count* (1x 4 ml EDTA) |
| **→ DISPENSE**<br>*Diary and home airflow monitoring device* | | Explain the use of Diary and compliance<br>Explain the use of home Airflow Monitoring device |

| | | |
|---|---|---|
| **REMARKS** | | |
| **Register in eCRF only when subject is eligible for baseline visit** | | **TRIAL URL**: https://www.data.castoredc.com/#studies |

**VISIT 4 – 23 (Days 2-46)**

| ITEM | Initials | Data / Comments |
|---|---|---|
| Visit date / number: | | _ _ / _ _ _ / _ _ _ _ ; _ _ _ |
| **REVIEW (S)AE** | | Change in (S)AE's since last visit? Yes / No<br>If yes, record on (S)AE form |
| **REVIEW CONCOMITANT MEDICATION** | | Change in medication since last visit? Yes / No<br>If yes, record on Medication form |
| **IN-/EXCLUSION CRITERIA**<br>*Follow protocol section 4.2/4.3* | | Complete checklist on In-/Exclusion criteria form |
| **PULMONARY/NASAL EXAMINATION** | | Status of body system? **N**ormal / **A**bnormal<br>Check body system and, if **A**bnormal, describe abnormality below<br>**N / A**<br>Nose □ / □ ………………………………………………………………………<br>Lung □ / □ ……………………………………………………………………… |
| **VITAL SIGNS**<br>*After at least 5 min in a semi-recumbent position* | . . : . . | Temperature: _ _ , _°C (tympanic) Respiratory rate: _ _ breaths/min<br>Pulse: _ _ _ beats/min Blood pressure (Sys/Dias): _ _ _ / _ _ _ mmHg |
| **FOT**<br>(see also prints out) | . . : . . | Resistance at 5Hz: _ _ _ _ cmH2O/L*s<br>Reactance at 5Hz: _ _ _ _ cmH2O/L*s<br>Resistance at 11Hz: _ _ _ _ cmH2O/L*s<br>Reactance at 11Hz: _ _ _ _ cmH2O/L*s<br>Resistance at 19Hz: _ _ _ _ cmH2O/L*s<br>Reactance at 19Hz: _ _ _ _ cmH2O/L*s<br>R5 – R19: _ _ _ _ cmH2O/L*s |
| **FeNO** | . . : . . | Has subject refrained from eating, drinking and strenuous exercise for at least 1 hr and consumed green vegetables within 24 hrs? Yes / No<br>_ _ _ ppb |

| | | |
|---|---|---|
| **VOCs_ GS-MS**<br>(see alsoTube Identification Form) | <br><br>. . : . .<br><br>. . : . .<br><br>. . : . . | Has subject refrained from eating, drinking and taking inhalation medication for at least 2 hours? Yes / No<br><br>Start 5 min tidal breathing<br><br>Collection of exhaled air in Tedlar bag<br><br>Adsorption of exhaled air sample on Tenax tubes |
| **VOCs_ SpiroNose**<br>(see also prints out Form) | <br><br><br><br>. . : . . | Did the subject rinse the mouth 3 times with water? Yes / No<br><br>Has the subject taken alcohol 12 hours before visit to the clinic?<br><br>Yes/No |
| **Exhaled Breath Condensate**<br>(see prints out for lab results) | . . : . . | Did the subject rinse the mouth before the exam? |
| **NASAL LAVAGE**<br>*(see Virology print outs for lab results)* | <br><br>. . : . . | _ _ , _ ml obtained after instillation of 8 ml 0.9% NaCl in R / L nostril<br><br>☐ Not required for this visit |
| **NASAL SWABS**<br>*(always check NOT to select the nostril used for the nasal lavage)* | . . : . . | ☐ Collected in R / L nostril<br>☐ Not required |

| | | |
|---|---|---|
| **SPUTUM INDUCTION**<br>*Perform FEV1 measurements post salbutamol and after each nebulisation period* | . . : . .<br><br>. . : . .<br><br>. . : . .<br><br>. . : . .<br><br>. . : . .<br><br>. . : . .<br><br>. . : . .<br><br>. . : . . | Post-salbutamol FEV1 (L): _ , _ _ (90% FEV1(L): _ , _ _ ) (80% FEV1(L): _ , _ _ )<br><br>$O_2$ Saturation (pre-induction): _ _ _ %<br><br>Post-bronchodilator FEV1 <50%predicted / ≥50% predicted *****<br>Post-bronchodilator FEV1 <1.0 L / 1.0 – 1.5 L / ≥ 1.5 L *****<br><br>Start try-out 2 min nebulisation period with 0.9% NaCl<br><br>FEV1 (L): _ , _ _ Fall ≤10% / 10-20% / >20%? * Sputum? Yes / No<br><br>Start 1st 5 min nebulisation period with 0.9% / 3% / 4.5% NaCl<br><br>FEV1 (L): _ , _ _ Fall ≤10% / 10-20% / >20%? * Sputum? Yes / No<br><br>Start 2nd 5 min nebulisation period with 0.9% / 3% / 4.5% NaCl<br><br>FEV1 (L): _ , _ _ Fall ≤10% / 10-20% / >20%? * Sputum? Yes / No<br><br>Start 3rd 5 min nebulisation period with 0.9% / 3% / 4.5% NaCl<br><br>FEV1 (L): _ , _ _ Fall ≤10% / 10-20% / >20%? * Sputum? Yes / No<br><br>Start 4th 5 min nebulisation period with 0.9% / 3% / 4.5% NaCl<br><br>FEV1 (L): _ , _ _ Fall ≤10% / 10-20% / >20%? * Sputum? Yes / No<br><br>Total time of induction: _ _ min<br><br>$O_2$ Saturation (post-induction): _ _ _ %<br><br>* select saline concentration to be used based on (changes in) FEV1<br><br>☐ Not required for this visit |
| **URINESAMPLING**<br>*(see print outs for lab results)* | . . : . . | ☐ Urine sample collected |
| **BLOODSAMPLING**<br>*(see print out for CBC lab result)* | . . : . . | Has subject fasted for at least 6 hours? Yes / No<br><br>☐ *Blood for Proteomics/Lipidomics/Metabolomics and hRV16 titre* (1x 10 ml Serum)<br><br>☐ *Blood for Standard Complete Blood Count* (1x 4 ml EDTA)<br><br>☐ Not required for this visit |
| **← COLLECT**<br>*Data home airflow monitoring device* | | Download data home airflow monitoring device. Sign and date print out of downloaded data |

| | | |
|---|---|---|
| ← **CHECK** | | Check compliance Diary and PEF/FEV1 values from home airflow monitoring device and retrain subject to use diary if necessary |
| → **DISPENSE**<br>*Diary* | | Retrain subject to use the diary if necessary |
| **REMARKS** | | |
| **Register in eCRF only when subject is eligible for baseline visit** | | **TRIAL URL**: https://www.data.castoredc.com/#studies |

**VISIT 24 (RV16-challenge) (Day 24)**

| ITEM | Initials | Data / Comments |
|---|---|---|
| Visit date: | | _ _ / _ _ _ / _ _ _ _ |
| **REVIEW (S)AE** | | Change in (S)AE's since last visit? Yes / No<br>If yes, record on (S)AE form |
| **REVIEW CONCOMITANT MEDICATION** | | Change in medication since last visit? Yes / No<br>If yes, record on Medication form |
| **IN-/EXCLUSION CRITERIA**<br>*Follow protocol section 4.2/4.3* | | Complete checklist on In-/Exclusion criteria form |
| **URINE PREGNANCY TEST**<br>*(see copy of result test)* | | If female with child bearing potential, perform urine pregnancy test<br>☐ Not applicable<br>☐ Positive<br>☐ Negative |
| **PULMONARY/NASAL EXAMINATION** | | Status of body system? **N**ormal / **A**bnormal<br>Check body system and, if **A**bnormal, describe abnormality below<br>**N / A**<br>Nose □ / □ ........................................<br>Lung □ / □ ........................................ |
| **VITAL SIGNS pre-challenge**<br>*After at least 5 min in a semi-recumbent position* | . . : . . | Temperature: _ _ , _°C (tympanic) Respiratory rate: _ _ breaths/min<br>Pulse: _ _ _ beats/min Blood pressure (Sys/Dias): _ _ _ / _ _ _ mmHg<br>$O_2$ Saturation: _ _ _ % |
| **FOT**<br>(see also prints out) | . . : . . | Resistance at 5Hz: _ _ _ _ cmH2O/L*s<br>Reactance at 5Hz: _ _ _ _ cmH2O/L*s<br>Resistance at 11Hz: _ _ _ _ cmH2O/L*s<br>Reactance at 11Hz: _ _ _ _ cmH2O/L*s<br>Resistance at 19Hz: _ _ _ _ cmH2O/L*s<br>Reactance at 19Hz: _ _ _ _ cmH2O/L*s<br>R5 – R19: _ _ _ _ cmH2O/L*s |

| | | |
|---|---|---|
| **FeNO** | . . : . . | Has subject refrained from eating, drinking and strenuous exercise for at least 1 hr and consumed green vegetables within 24 hrs? Yes / No<br>_ _ _ ppb |
| **VOCs_ GS-MS**<br>(see alsoTube Identification Form) | <br>. . : . .<br>. . : . .<br>. . : . . | Has subject refrained from eating, drinking and taking inhalation medication for at least 2 hours? Yes / No<br>Start 5 min tidal breathing<br>Collection of exhaled air in Tedlar bag<br>Adsorption of exhaled air sample on Tenax tubes |
| **VOCs_ SpiroNose**<br>(see also prints out Form) | . . : . . | Did the subject rinse the mouth 3 times with water? Yes / No<br>Has the subject taken alcohol 12 hours before visit to the clinic?<br>Yes/No |
| **NASAL LAVAGE**<br>*(see Virology print outs for lab results)* | . . : . . | _ _ , _ ml obtained after instillation of 8 ml 0.9% NaCl in R / L nostril |
| **NASAL SWABS**<br>*(always check NOT to select the nostril used for the nasal lavage)* | . . : . . | ☐ Collected in R / L nostril |
| **Exhaled Breath Condensate**<br>(see prints out for lab results) | . . : . . | Did the subject rinse the mouth before the exam? |

| | | |
|---|---|---|
| **SPUTUM INDUCTION**<br>*Perform FEV1 measurements post salbutamol and after each nebulisation period* | . . : . .<br><br>. . : . .<br><br><br>. . : . .<br><br>. . : . .<br><br>. . : . .<br><br>. . : . .<br><br>. . : . .<br><br><br>. . : . . | Post-salbutamol FEV1 (L): _ , _ _ (90% FEV1(L): _ , _ _ ) (80% FEV1(L): _ , _ _ )<br><br>$O_2$ Saturation (pre-induction): _ _ _ %<br><br>Post-bronchodilator FEV1 <50%predicted / ≥50% predicted *<br>Post-bronchodilator FEV1 <1.0 L / 1.0 – 1.5 L / ≥ 1.5 L *<br><br>Start try-out 2 min nebulisation period with 0.9% NaCl<br><br>FEV1 (L): _ , _ _ Fall ≤10% / 10-20% / >20%? * Sputum? Yes / No<br><br>Start 1st 5 min nebulisation period with 0.9% / 3% / 4.5% NaCl<br><br>FEV1 (L): _ , _ _ Fall ≤10% / 10-20% / >20%? * Sputum? Yes / No<br><br>Start 2nd 5 min nebulisation period with 0.9% / 3% / 4.5% NaCl<br><br>FEV1 (L): _ , _ _ Fall ≤10% / 10-20% / >20%? * Sputum? Yes / No<br><br>Start 3rd 5 min nebulisation period with 0.9% / 3% / 4.5% NaCl<br><br>FEV1 (L): _ , _ _ Fall ≤10% / 10-20% / >20%? * Sputum? Yes / No<br><br>Start 4th 5 min nebulisation period with 0.9% / 3% / 4.5% NaCl<br><br>FEV1 (L): _ , _ _ Fall ≤10% / 10-20% / >20%? * Sputum? Yes / No<br><br>Total time of induction: _ _ min<br><br>$O_2$ Saturation (post-induction): _ _ _ %<br><br>* select saline concentration to be used based on (changes in) FEV1<br><br>☐ Not required for this visit |
| **URINESAMPLING**<br>*(see print outs for lab results)* | . . : . . | ☐ Urine sample collected |
| **BLOODSAMPLING**<br>*(see print out for CBC lab result)* | . . : . . | Has subject fasted for at least 6 hours? Yes / No<br><br>☐ *Blood for Proteomics/Lipidomics/Metabolomics and hRV16 titre* (1x 10 ml Serum)<br>☐ *Blood for Standard Complete Blood Count* (1x 4 ml EDTA)<br><br>☐ Not required for this visit |
| **hRV16 inoculation**<br>*(see also dilution form)* | . . : . . | Is hRV16 (dose 100 TCID50) administered? Yes / No<br><br>If Yes, which nostril? Left / Right |

| | | |
|---|---|---|
| **VITAL SIGNS 5 min post-challenge**<br>*After at least 5 min in a semi-recumbent position* | . . : . . | Temperature: _ _ , _°C (tympanic) Respiratory rate: _ _ breaths/min<br>Pulse: _ _ _ beats/min Blood pressure (Sys/Dias): _ _ _ / _ _ _ mmHg<br>$O_2$ Saturation: _ _ _ % |
| ← **COLLECT**<br>*Data home airflow monitoring device* | | Download data home airflow monitoring device. Sign and date print out of downloaded data |
| ← **CHECK** | | Check compliance Diary and PEF/FEV1 values from home airflow monitoring device and retrain subject to use diary if necessary |
| → **DISPENSE**<br>*Diary* | | Retrain subject to use the diary if necessary |
| **REMARKS** | | |

| | | |
|---|---|---|
| **EVENTS OF CLINICAL INTEREST**<br>(see protocol section 5) | | Did an Event of Clinical Interest occur? Yes / No<br>If Yes, check event below<br>☐ Overdose of virus<br>☐ Systemic steroids requirement to treat asthma exacerbation related to virus challenge<br>☐ Acute reaction to virus challenge as defined below when confirmed through repeat measurement and considered potentially associated with administration of the virus (see grading scales in protocol section 12.4)<br>☐ One of these vital sign findings within 4 h of challenge:<br>* Fever (≥ 38.5 °C)<br>*Tachycardia (hr>115/min)<br>* Bradycardia (hr<50/min, unless within 5 bpm of baseline, and/or asymptomatic)<br>* Hypertension (sbp>155, dbp>100)<br>* Hypotension (sbp<90, unless within 3 mm Hg of resting sbp)<br>* Tachypnea (rate >25/min)<br>* $SpO_2$ (<92%, or >4% decrease from baseline)<br>☐ >20% decrease in FEV1 (within 4 h of challenge) relative to baseline<br>☐ Dyspnea associated with drop in FEV1 (within 4 h of challenge) that is unresponsive to a bronchodilator rescue agent within 20 minutes<br>☐ One of these symptom findings within 24h of challenge:<br>* Nausea/Vomiting (grade 2+)<br>* Diarrhea (grade 3+)<br>* Headache (grade 3+)<br>* Fatigue (grade 2+)<br>* Myalgia (grade 2+)<br>☐ Grade 2+ deviation from normal values of liver-related laboratory parameters at any time between challenge and day 14 (see table in protocol section 12.4) |
| **Register in eCRF only when subject is eligible for baseline visit** | | **TRIAL URL**: https://www.data.castoredc.com/#studies |

**VISIT 25-35 (DAYS 51-74)**

| ITEM | Initials | Data / Comments |
|---|---|---|
| Visit date / number: | | _ _ / _ _ _ / _ _ _ _ ; _ _ _ |
| **REVIEW (S)AE** | | Change in (S)AE's since last visit? Yes / No<br>If yes, record on (S)AE form |
| **REVIEW CONCOMITANT MEDICATION** | | Change in medication since last visit? Yes / No<br>If yes, record on Medication form |
| **IN-/EXCLUSION CRITERIA**<br>*Follow protocol section 4.2/4.3* | | Complete checklist on In-/Exclusion criteria form |
| **PULMONARY/NASAL EXAMINATION** | | Status of body system? **N**ormal / **A**bnormal<br>Check body system and, if **A**bnormal, describe abnormality below<br>**N / A**<br>Nose □ / □ ......................................................<br>Lung □ / □ ...................................................... |
| **VITAL SIGNS**<br>*After at least 5 min in a semi-recumbent position* | . . : . . | Temperature: _ _ , _°C (tympanic) Respiratory rate: _ _ breaths/min<br>Pulse: _ _ _ beats/min Blood pressure (Sys/Dias): _ _ _ / _ _ _ mmHg |
| **FOT**<br>(see also prints out) | . . : . . | Resistance at 5Hz: _ _ _ _ cmH2O/L*s<br>Reactance at 5Hz: _ _ _ _ cmH2O/L*s<br>Resistance at 11Hz: _ _ _ _ cmH2O/L*s<br>Reactance at 11Hz: _ _ _ _ cmH2O/L*s<br>Resistance at 19Hz: _ _ _ _ cmH2O/L*s<br>Reactance at 19Hz: _ _ _ _ cmH2O/L*s<br>R5 – R19: _ _ _ _ cmH2O/L*s |
| **FeNO** | . . : . . | Has subject refrained from eating, drinking and strenuous exercise for at least 1 hr and consumed green vegetables within 24 hrs? Yes / No<br>_ _ _ ppb |

| | | |
|---|---|---|
| **VOCs_ GS-MS**<br>(see alsoTube Identification Form) | <br><br>. . : . .<br>. . : . .<br>. . : . . | Has subject refrained from eating, drinking and taking inhalation medication for at least 2 hours? Yes / No<br>Start 5 min tidal breathing<br>Collection of exhaled air in Tedlar bag<br>Adsorption of exhaled air sample on Tenax tubes |
| **VOCs_ SpiroNose**<br>(see also prints out Form) | . . : . . | Did the subject rinse the mouth 3 times with water? Yes / No<br>Has the subject taken alcohol 12 hours before visit to the clinic?<br>Yes/No |
| **NASAL LAVAGE**<br>*(see Virology print outs for lab results)* | . . : . . | _ _ , _ ml obtained after instillation of 8 ml 0.9% NaCl in R / L nostril<br>☐ Not required for this visit |
| **NASAL SWABS**<br>*(always check NOT to select the nostril used for the nasal lavage)* | . . : . . | ☐ Collected in R / L nostril<br>☐ Not required |
| **Exhaled Breath Condensate**<br>(see prints out for lab results) | . . : . . | Did the subject rinse the mouth before the exam? |

| | | |
|---|---|---|
| **SPUTUM INDUCTION**<br>*Perform FEV1 measurements post salbutamol and after each nebulisation period* | . . : . .<br><br>. . : . .<br><br>. . : . .<br><br>. . : . .<br><br>. . : . .<br><br>. . : . .<br><br>. . : . .<br><br>. . : . . | Post-salbutamol FEV1 (L): _ , _ _ (90% FEV1(L): _ , _ _ ) (80% FEV1(L): _ , _ _ )<br><br>$O_2$ Saturation (pre-induction): _ _ _ %<br><br>Post-bronchodilator FEV1 <50%predicted / ≥50% predicted *<br>Post-bronchodilator FEV1 <1.0 L / 1.0 – 1.5 L / ≥ 1.5 L *<br><br>Start try-out 2 min nebulisation period with 0.9% NaCl<br><br>FEV1 (L): _ , _ _ Fall ≤10% / 10-20% / >20%? * Sputum? Yes / No<br><br>Start 1st 5 min nebulisation period with 0.9% / 3% / 4.5% NaCl<br><br>FEV1 (L): _ , _ _ Fall ≤10% / 10-20% / >20%? * Sputum? Yes / No<br><br>Start 2nd 5 min nebulisation period with 0.9% / 3% / 4.5% NaCl<br><br>FEV1 (L): _ , _ _ Fall ≤10% / 10-20% / >20%? * Sputum? Yes / No<br><br>Start 3rd 5 min nebulisation period with 0.9% / 3% / 4.5% NaCl<br><br>FEV1 (L): _ , _ _ Fall ≤10% / 10-20% / >20%? * Sputum? Yes / No<br><br>Start 4th 5 min nebulisation period with 0.9% / 3% / 4.5% NaCl<br><br>FEV1 (L): _ , _ _ Fall ≤10% / 10-20% / >20%? * Sputum? Yes / No<br><br>Total time of induction: _ _ min<br><br>$O_2$ Saturation (post-induction): _ _ _ %<br><br>* select saline concentration to be used based on (changes in) FEV1<br><br>☐ Not required for this visit |
| **URINESAMPLING**<br>*(see print outs for lab results)* | . . : . . | ☐ Urine sample collected |
| **BLOODSAMPLING**<br>*(see print out for CBC lab result)* | . . : . . | Has subject fasted for at least 6 hours? Yes / No<br><br>☐ *Blood for Proteomics/Lipidomics/Metabolomics and hRV16 titre* (1x 10 ml Serum)<br>☐ *Blood for Standard Complete Blood Count* (1x 4 ml EDTA)<br><br>☐ Not required for this visit |
| **← COLLECT**<br>*Data home airflow monitoring device* | | Download data home airflow monitoring device. Sign and date print out of downloaded data |

| | | |
|---|---|---|
| ← **CHECK** | | Check compliance Diary and PEF/FEV1 values from home airflow monitoring device and retrain subject to use diary if necessary |
| → **DISPENSE**<br>*Diary* | | Retrain subject to use the diary if necessary |
| **EVENTS OF CLINICAL INTEREST**<br>*(see protocol section 5)* | | Did an Event of Clinical Interest occur? Yes / No<br>If Yes, check event below<br>☐ Systemic steroids requirement to treat asthma exacerbation related to virus challenge<br>☐ Acute reaction to virus challenge as defined below when confirmed through repeat measurement and considered potentially associated with administration of the virus (see grading scales in protocol section 12.4)<br>☐ One of these symptom findings within 24h of challenge:<br>* Nausea/Vomiting (grade 2+)<br>* Diarrhea (grade 3+)<br>* Headache (grade 3+)<br>* Fatigue (grade 2+)<br>* Myalgia (grade 2+)<br>☐ Grade 2+ deviation from normal values of liver-related laboratory parameters at any time between challenge and day 14 (see table in protocol section 12.4) |
| **REMARKS** | | |
| **Register in eCRF only when subject is eligible for baseline visit** | | **TRIAL URL**: https://www.data.castoredc.com/#studies |

**END OF STUDY**

Date of study termination: _ _ / _ _ _ / _ _ _ _          Date of last contact (visit to AMC): _ _ / _ _ _ / _ _ _ _

Was the study completed? Yes / No

If No, specify reason of discontinuation:

☐ Adverse Event

☐ Death

☐ Pregnancy

☐ Lack of efficacy/qualifying event, specify below

☐ Technical problems

☐ Protocol violation, specify below

☐ Withdrawal Consent, specify below

☐ Lost to follow-up, specify below

☐ Screen failure, specify below

☐ Principal investigator/physician decision, specify below

☐ Sponsor decision, specify below

☐ Other, specify below

……………………………………………………………………………………………………………………

……………………………………………………………………………………………………………………

……………………………………………………………………………………………………………………

Completed by (initials + date):

**DATA ENTRY CHECK FORM**

*'I hereby declare that all data written in this worksheet are complete and accurate'*

Name: ……………………………………………………………………………………………….

Position: ……………………………………………………………………………………………..

Signature: …………………………………………………………………………………………...

Date: _ _ / _ _ _ / _ _ _ _
