## Supplemental Figures S1 through S20 for "Can measurements of inflammatory biomarkers be used to spot viral infections?"

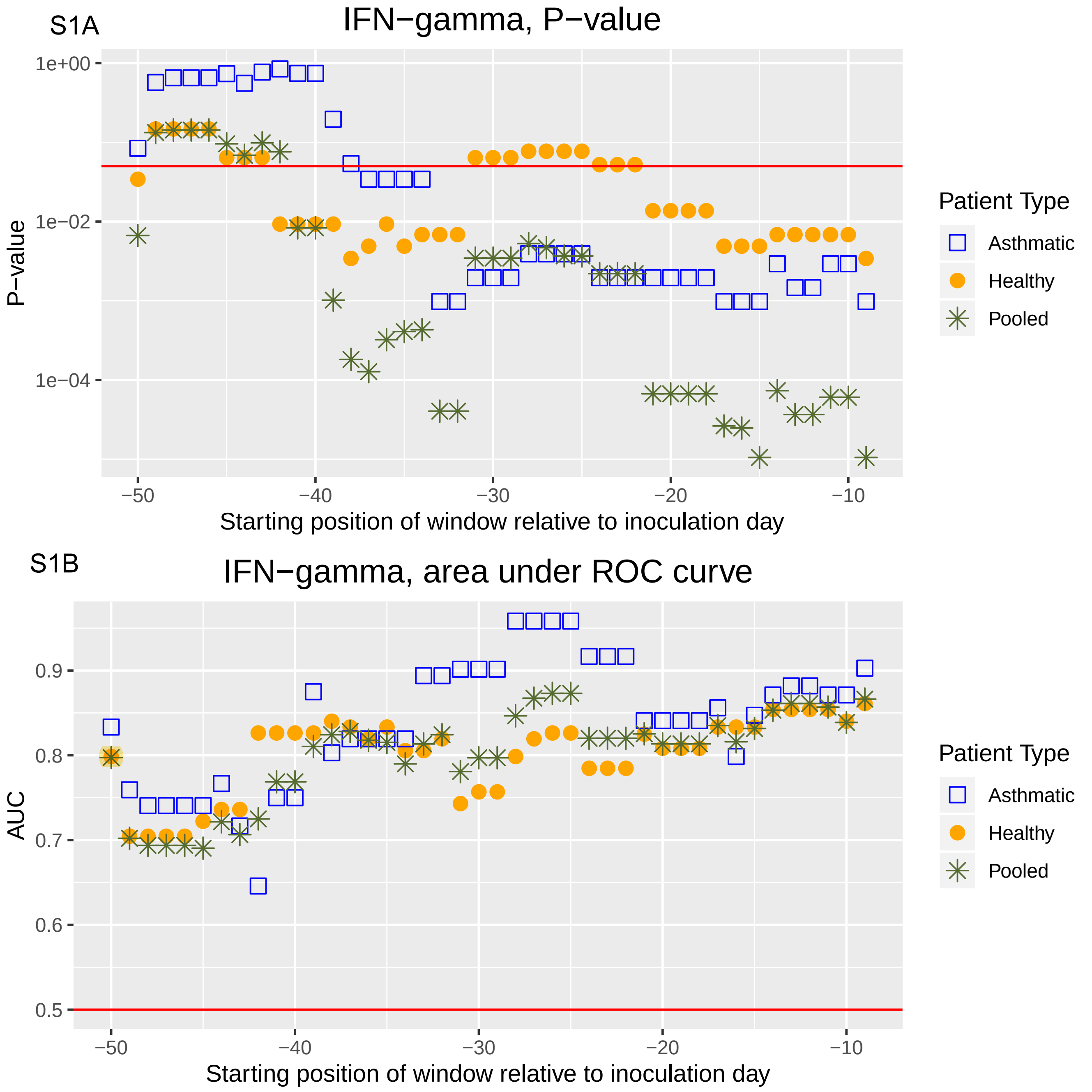

S1A
IFN−gamma, P−value
P−value
1e+00
1e−02
1e−04
−50
−40
−30
−20
−10
Starting position of window relative to inoculation day
Patient Type
Asthmatic
Healthy
Pooled
S1B
IFN−gamma, area under ROC curve
AUC
0.9
0.8
0.7
0.6
0.5
−50
−40
−30
−20
−10
Starting position of window relative to inoculation day
Patient Type
Asthmatic
Healthy
Pooled

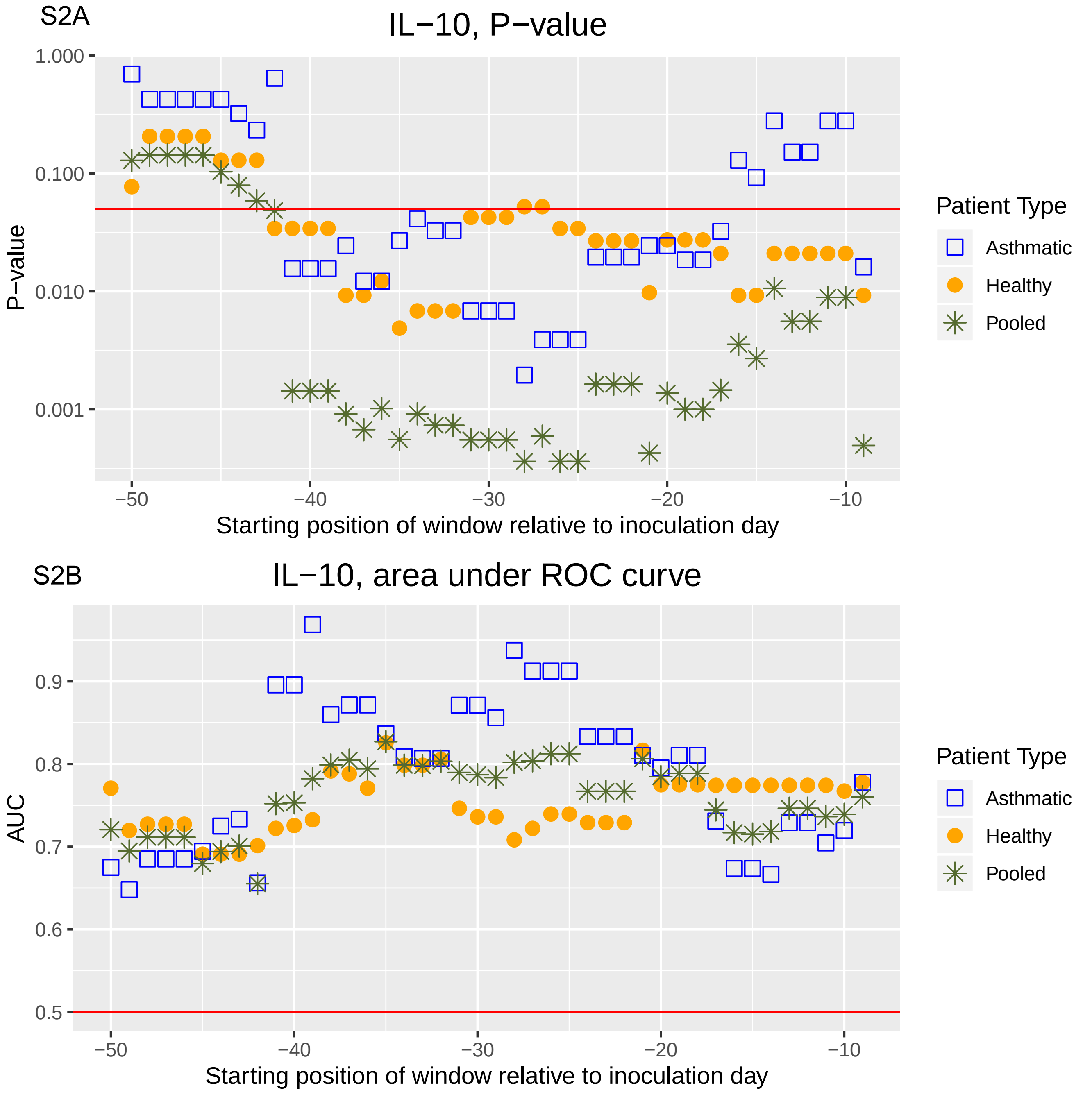
S2A
IL−10, P−value
P−value
1.000
0.100
0.010
0.001
−50
−40
−30
−20
−10
Starting position of window relative to inoculation day
Patient Type
Asthmatic
Healthy
Pooled
S2B
IL−10, area under ROC curve
AUC
0.9
0.8
0.7
0.6
0.5
−50
−40
−30
−20
−10
Starting position of window relative to inoculation day
Patient Type
Asthmatic
Healthy
Pooled

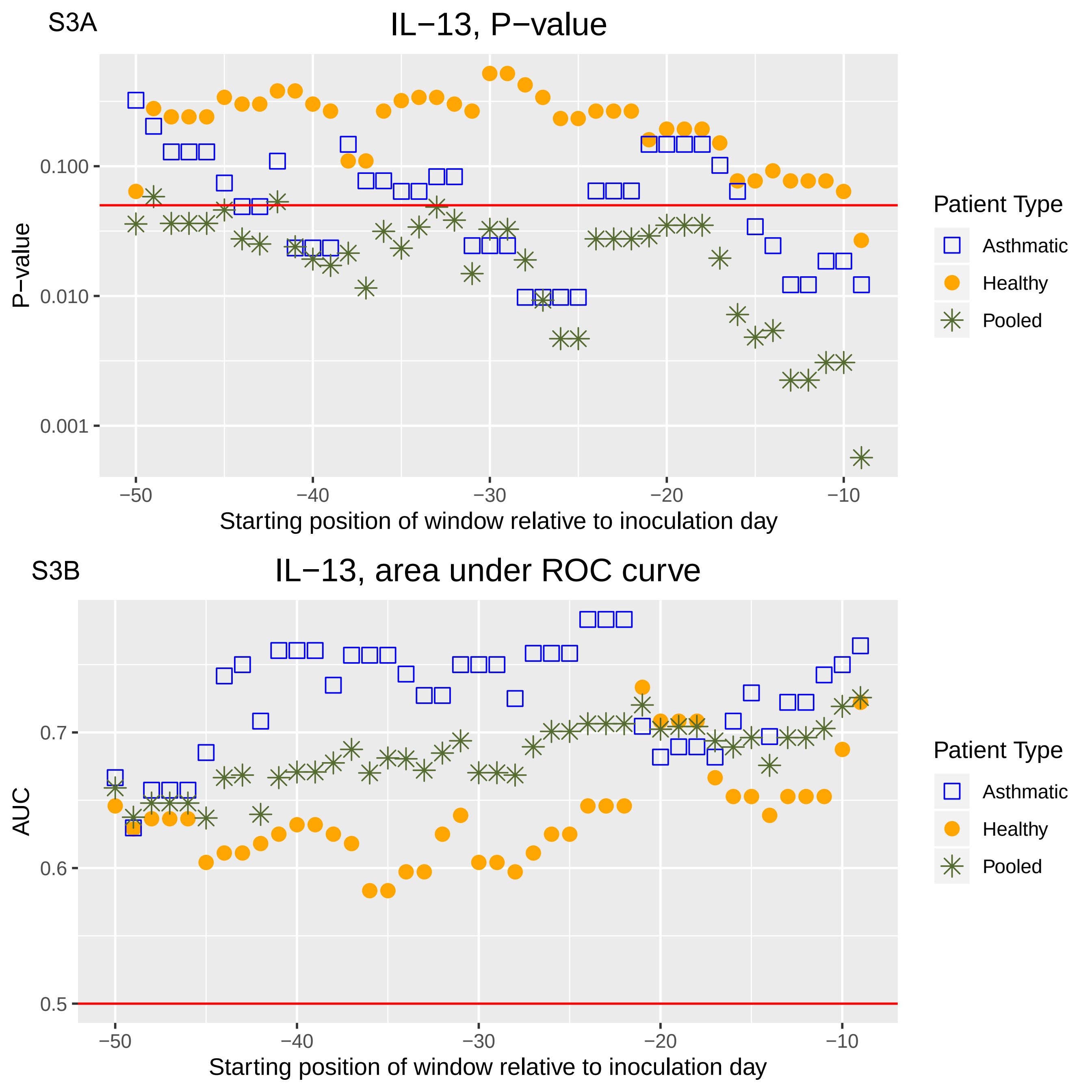
S3A
IL−13, P−value
P−value
0.100
0.010
0.001
−50
−40
−30
−20
−10
Starting position of window relative to inoculation day
Patient Type
Asthmatic
Healthy
Pooled
S3B
IL−13, area under ROC curve
AUC
0.7
0.6
0.5
−50
−40
−30
−20
−10
Starting position of window relative to inoculation day
Patient Type
Asthmatic
Healthy
Pooled

S4A

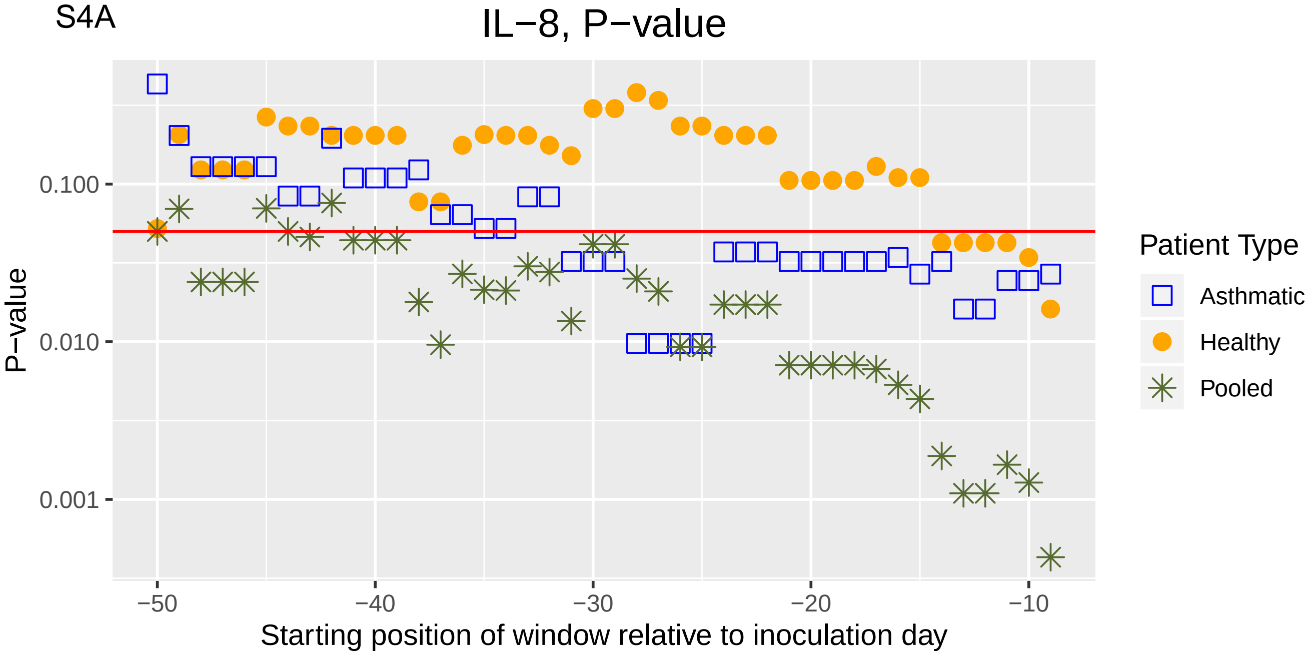
IL−8, P−value
P−value
0.100
0.010
0.001
−50
−40
−30
−20
−10
Starting position of window relative to inoculation day
Patient Type
Asthmatic
Healthy
Pooled

S4B

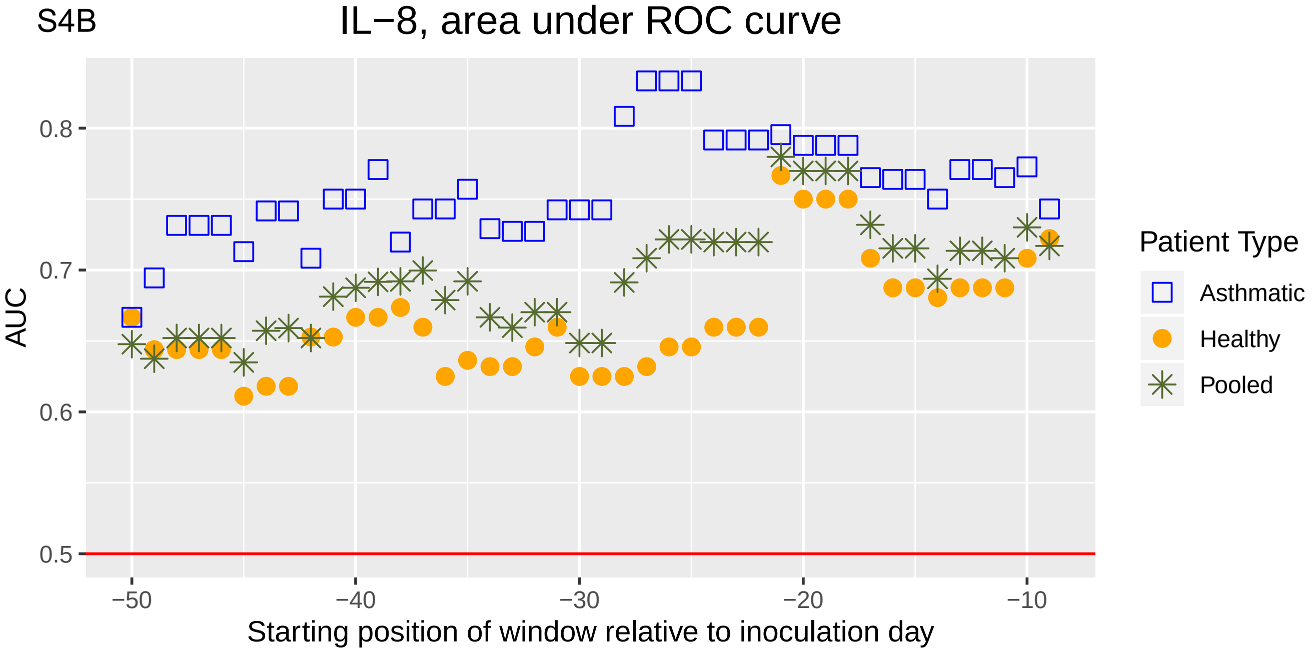
IL−8, area under ROC curve
AUC
0.8
0.7
0.6
0.5
−50
−40
−30
−20
−10
Starting position of window relative to inoculation day
Patient Type
Asthmatic
Healthy
Pooled

S5A

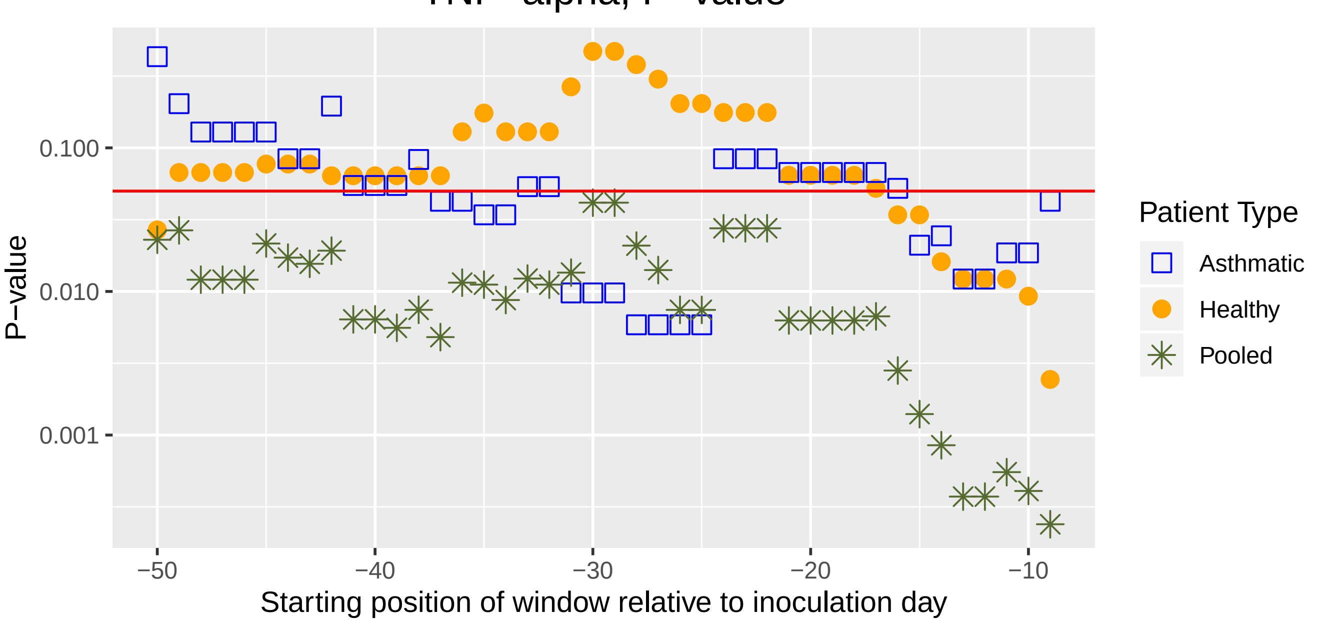
TNF−alpha, P−value
P−value
0.100
0.010
0.001
−50
−40
−30
−20
−10
Starting position of window relative to inoculation day
Patient Type
Asthmatic
Healthy
Pooled

S5B

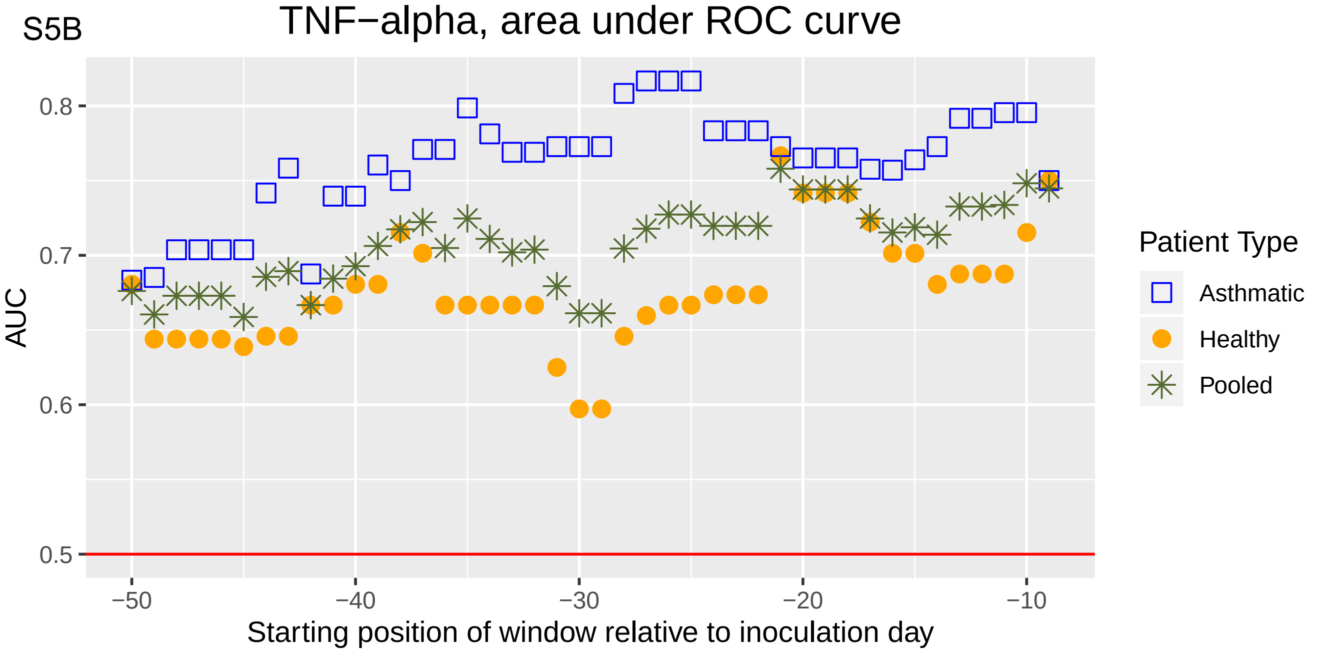
TNF−alpha, area under ROC curve
AUC
0.8
0.7
0.6
0.5
−50
−40
−30
−20
−10
Starting position of window relative to inoculation day
Patient Type
Asthmatic
Healthy
Pooled

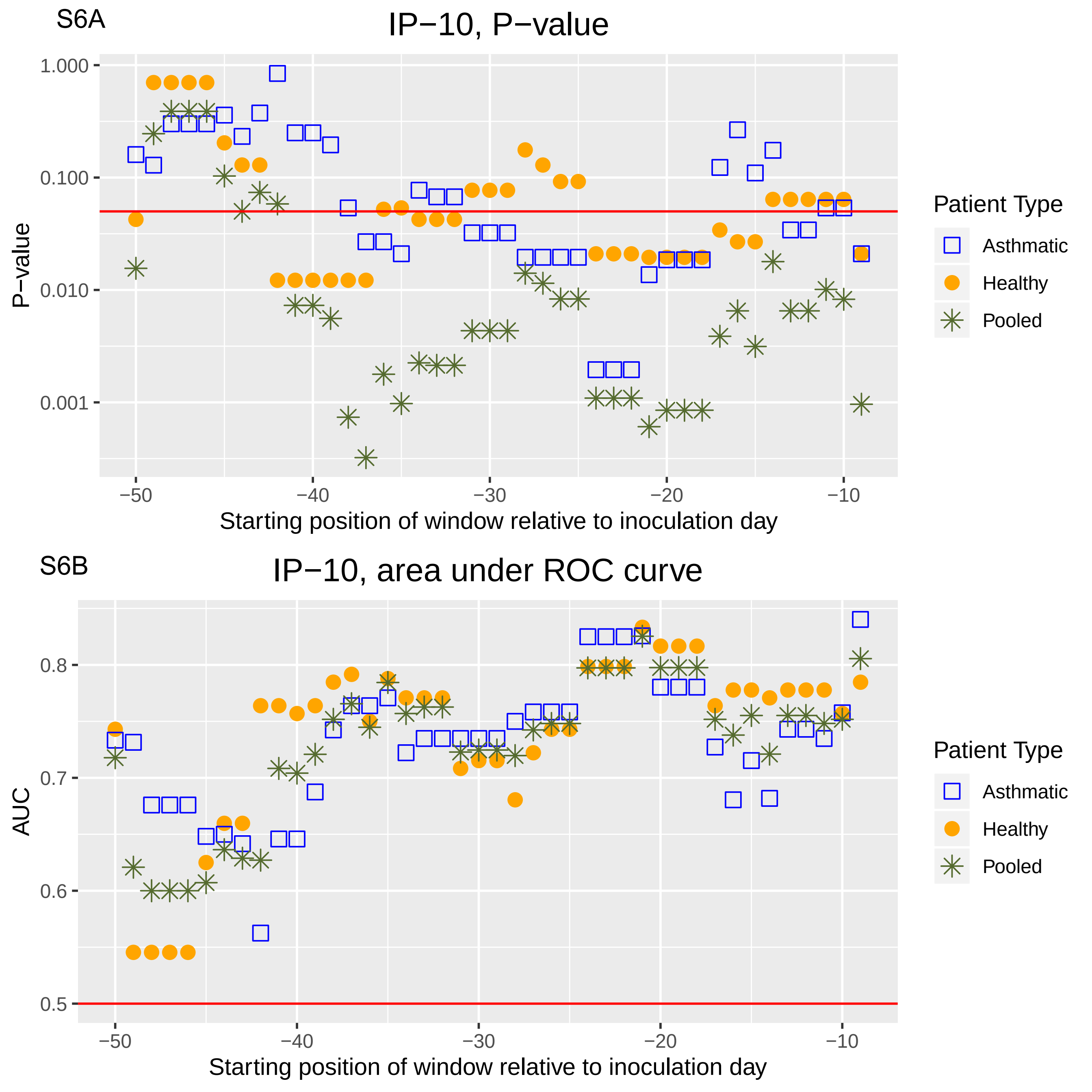

S6A
IP−10, P−value
P−value
1.000
0.100
0.010
0.001
−50
−40
−30
−20
−10
Starting position of window relative to inoculation day
Patient Type
Asthmatic
Healthy
Pooled
S6B
IP−10, area under ROC curve
AUC
0.8
0.7
0.6
0.5
−50
−40
−30
−20
−10
Starting position of window relative to inoculation day
Patient Type
Asthmatic
Healthy
Pooled

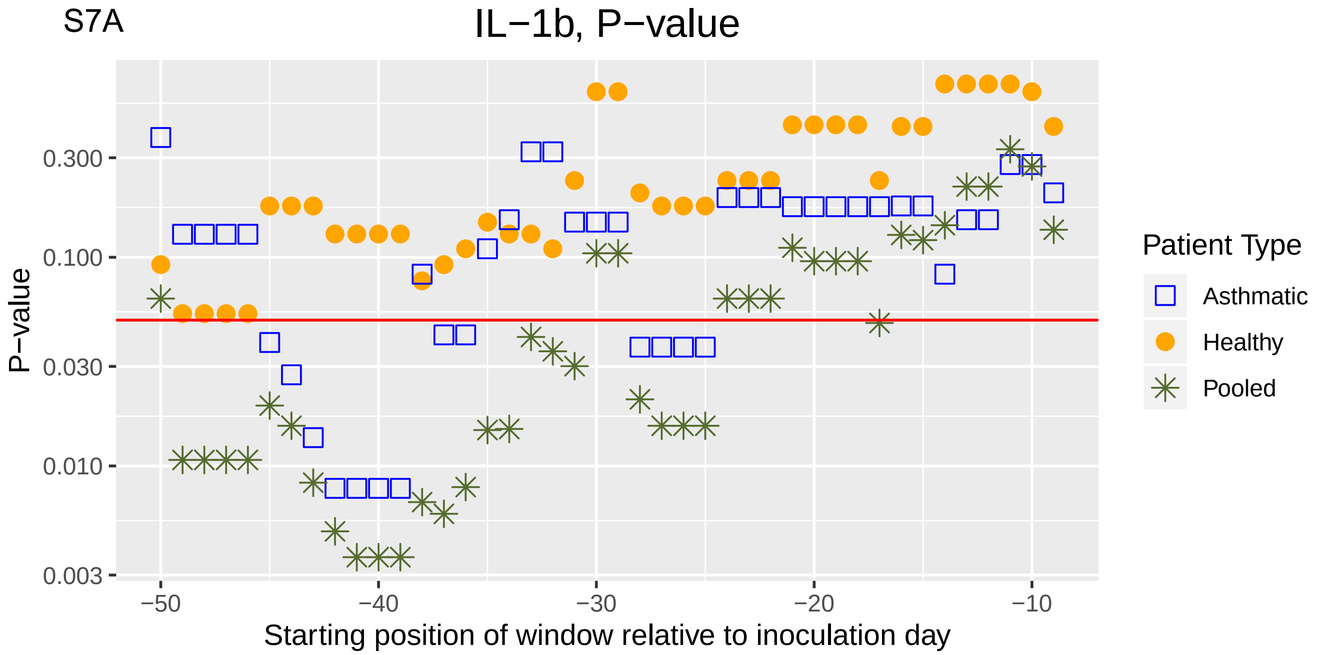
S7A
IL−1b, P−value
P−value
0.300
0.100
0.030
0.010
0.003
−50
−40
−30
−20
−10
Starting position of window relative to inoculation day
Patient Type
Asthmatic
Healthy
Pooled

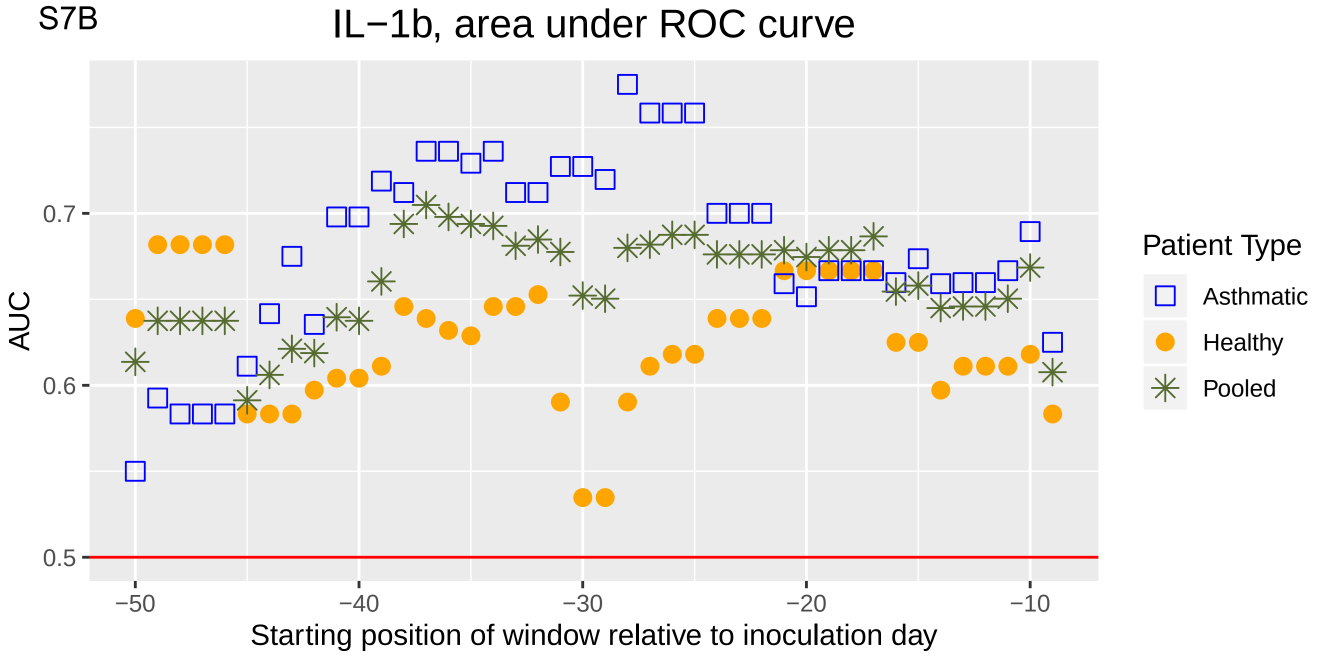
S7B
IL−1b, area under ROC curve
AUC
0.7
0.6
0.5
−50
−40
−30
−20
−10
Starting position of window relative to inoculation day
Patient Type
Asthmatic
Healthy
Pooled

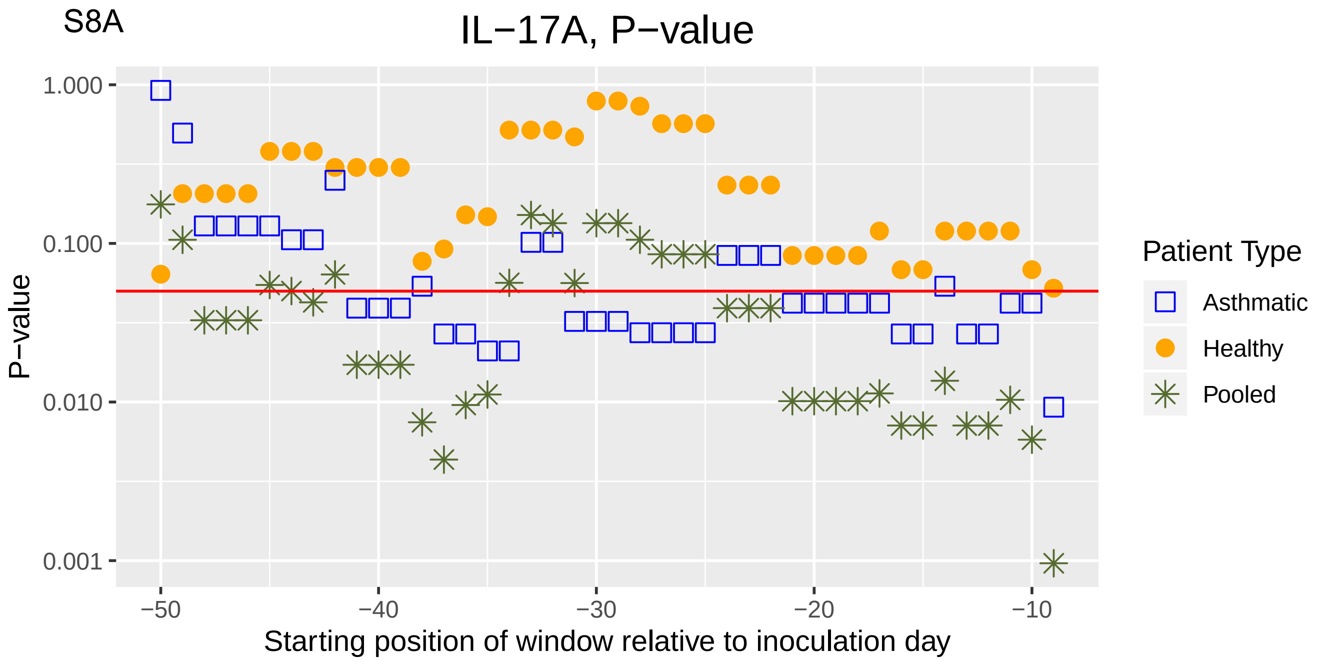
S8A
IL−17A, P−value
P−value
1.000
0.100
0.010
0.001
−50
−40
−30
−20
−10
Starting position of window relative to inoculation day
Patient Type
Asthmatic
Healthy
Pooled

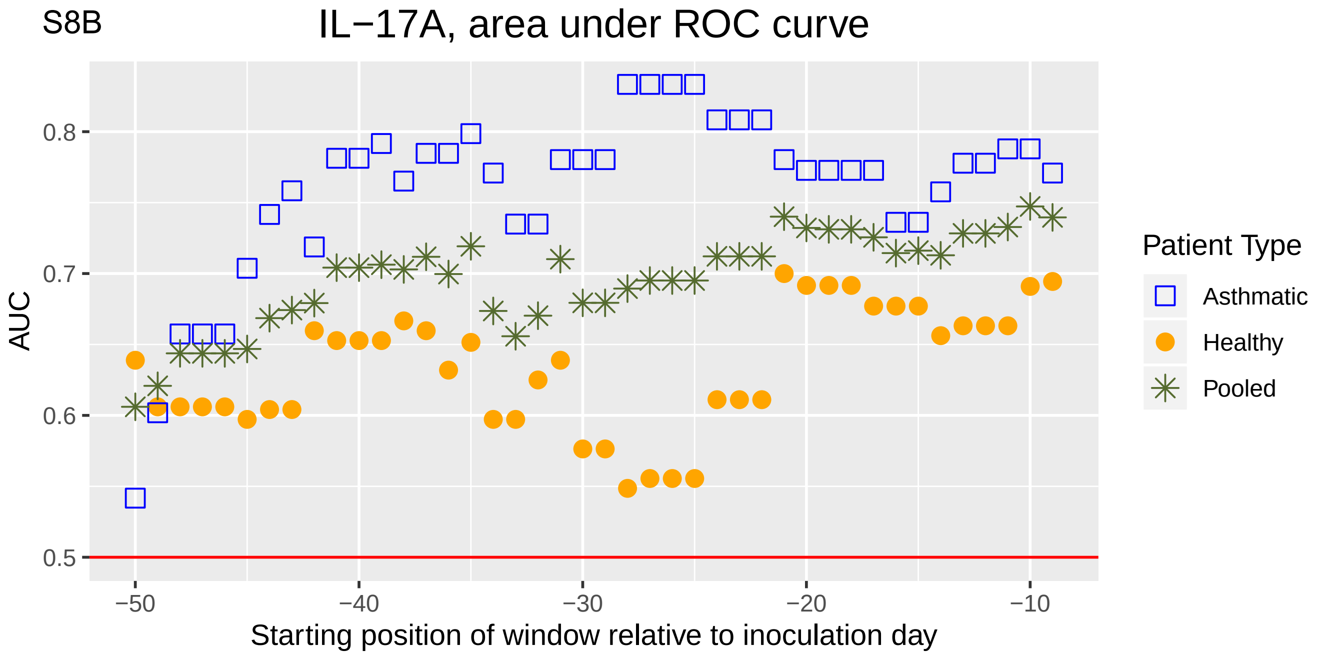
S8B
IL−17A, area under ROC curve
AUC
0.8
0.7
0.6
0.5
−50
−40
−30
−20
−10
Starting position of window relative to inoculation day
Patient Type
Asthmatic
Healthy
Pooled

S9A

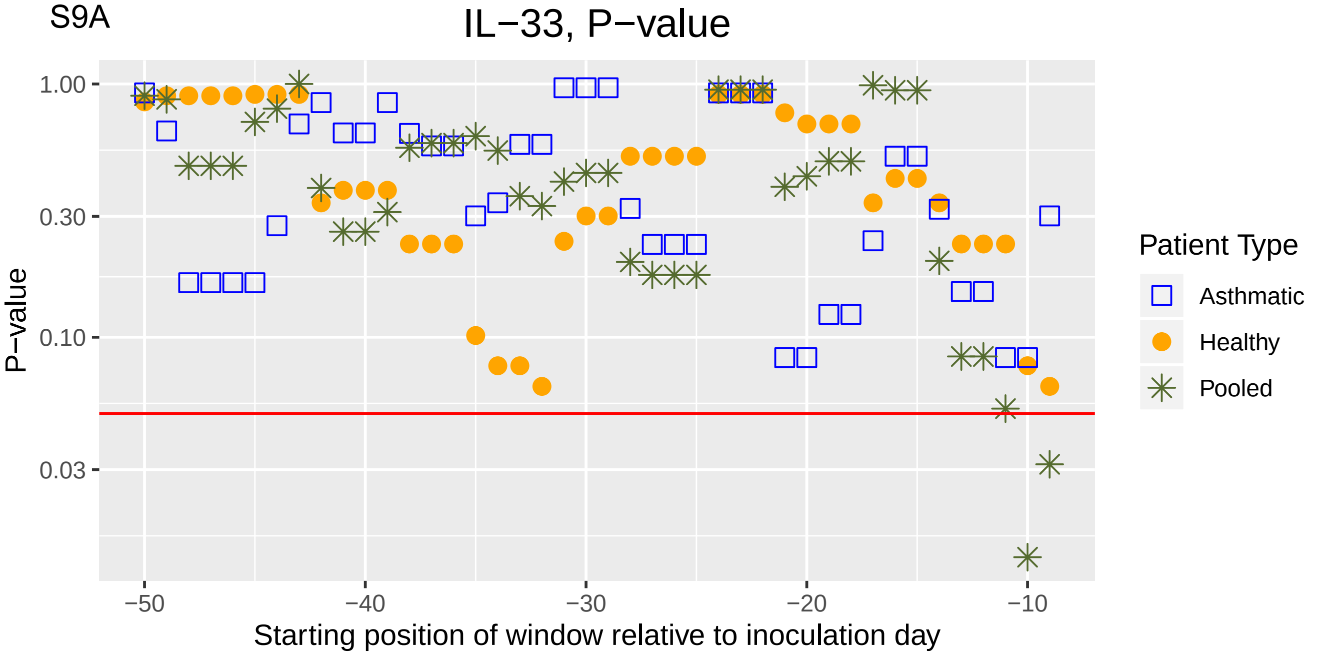
IL−33, P−value
P−value
1.00
0.30
0.10
0.03
−50
−40
−30
−20
−10
Starting position of window relative to inoculation day
Patient Type
Asthmatic
Healthy
Pooled

S9B

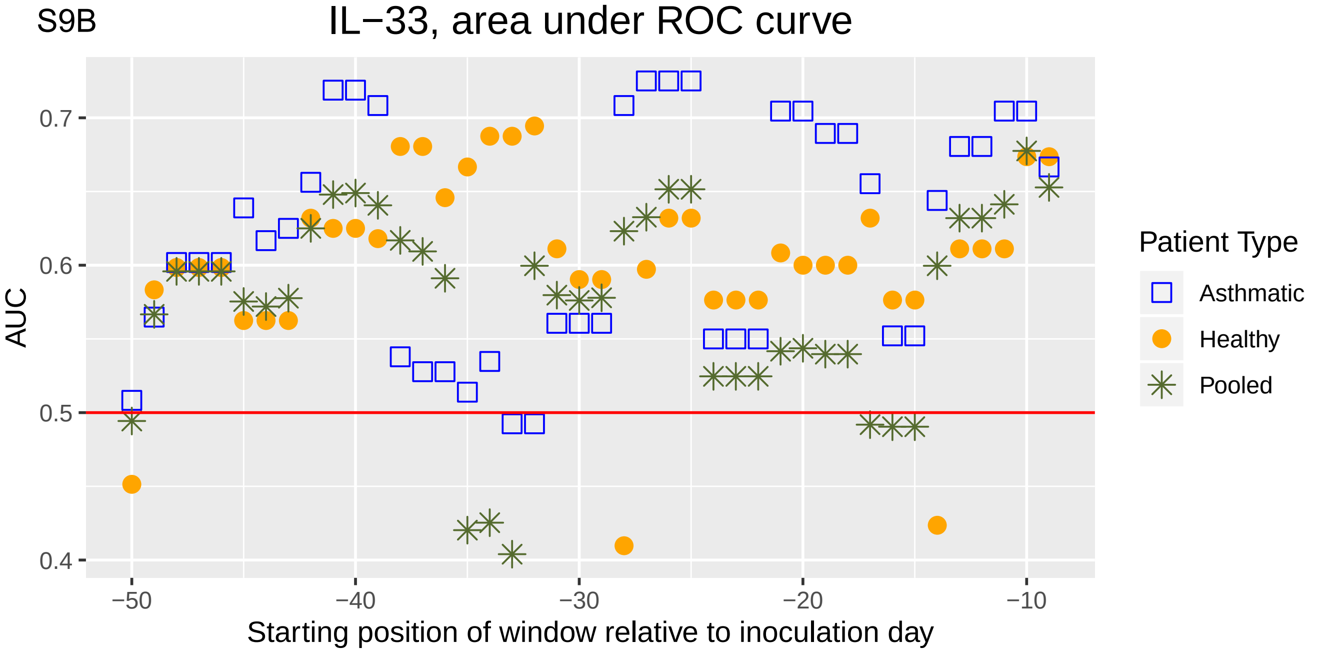
IL−33, area under ROC curve
AUC
0.7
0.6
0.5
0.4
−50
−40
−30
−20
−10
Starting position of window relative to inoculation day
Patient Type
Asthmatic
Healthy
Pooled

S10A

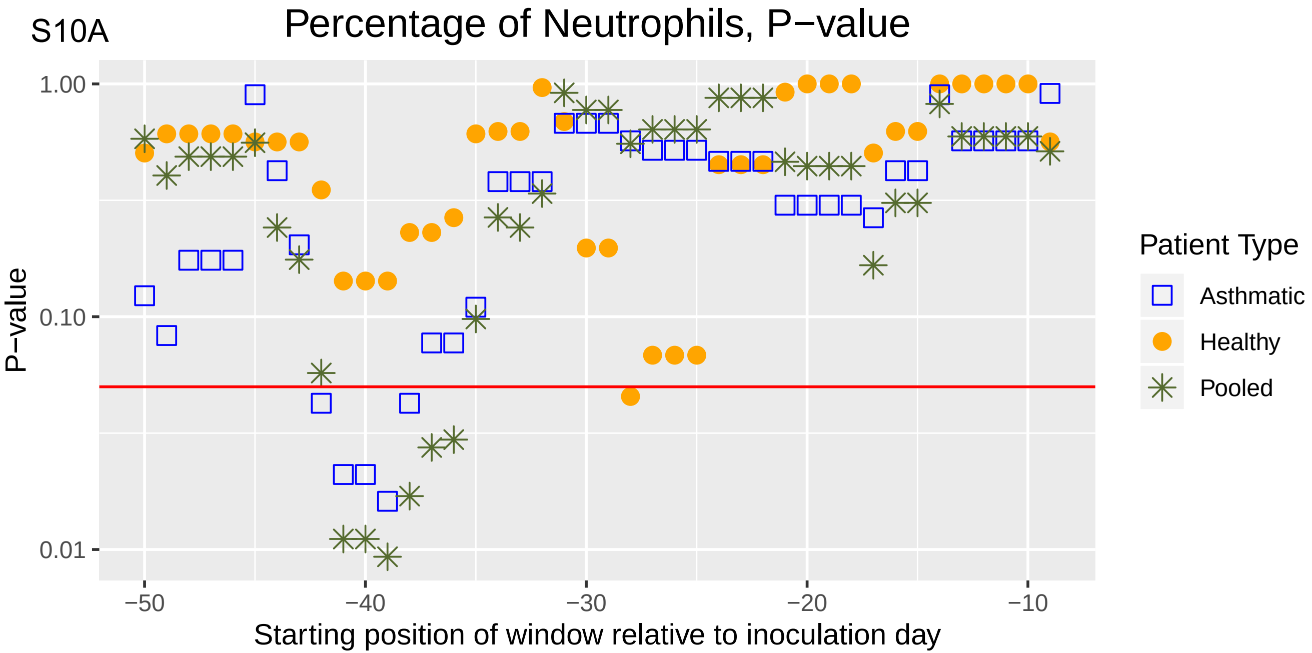
Percentage of Neutrophils, P−value
P−value
1.00
0.10
0.01
−50
−40
−30
−20
−10
Starting position of window relative to inoculation day
Patient Type
Asthmatic
Healthy
Pooled

S10B

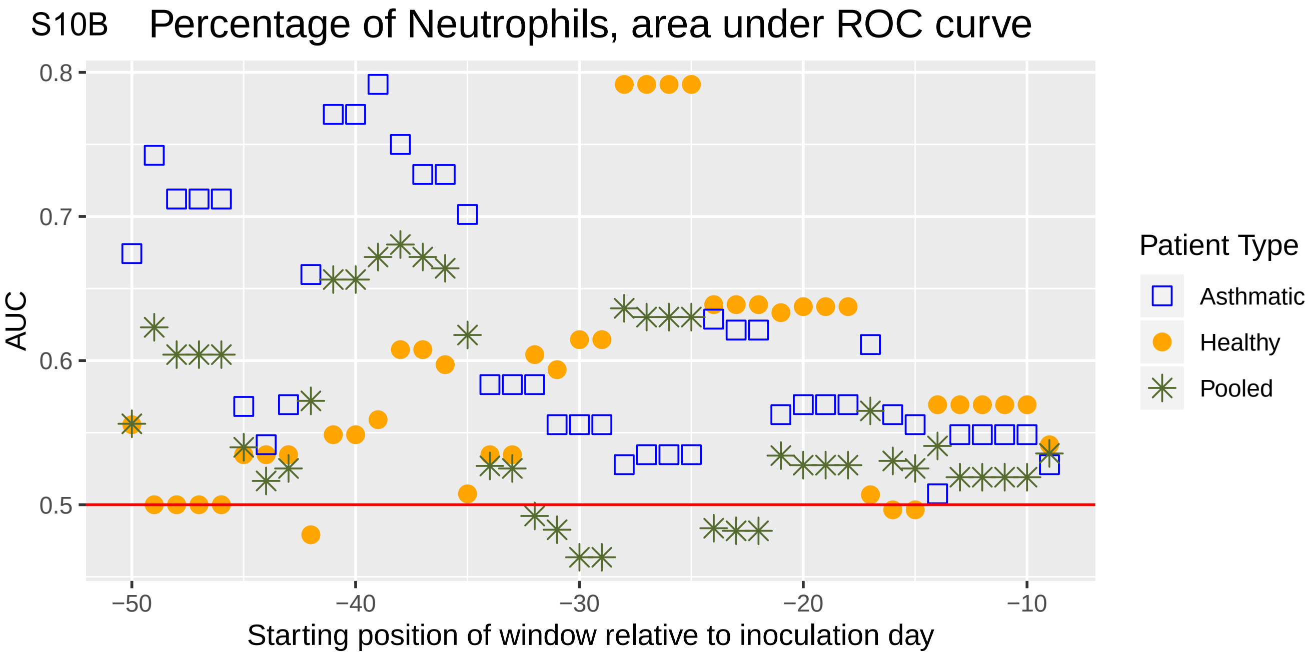
Percentage of Neutrophils, area under ROC curve
AUC
0.8
0.7
0.6
0.5
−50
−40
−30
−20
−10
Starting position of window relative to inoculation day
Patient Type
Asthmatic
Healthy
Pooled

S11A

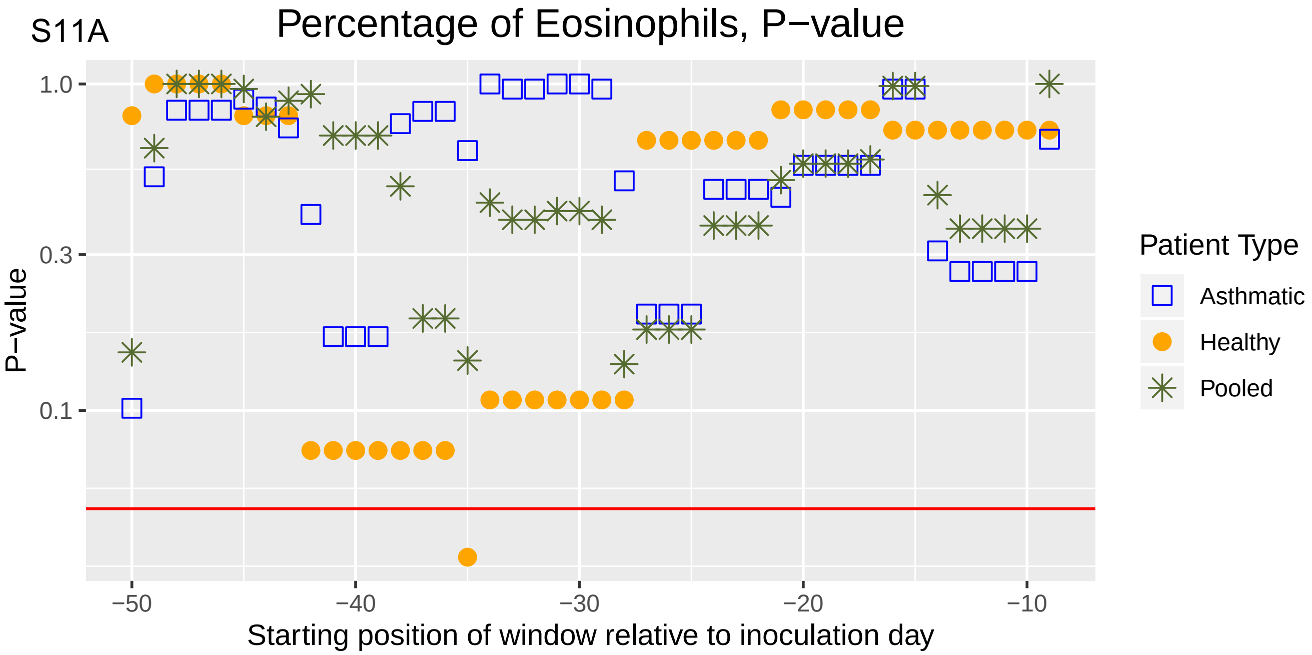

Percentage of Eosinophils, P−value
1.0
0.3
0.1
P−value
−50
−40
−30
−20
−10
Starting position of window relative to inoculation day
Patient Type
Asthmatic
Healthy
Pooled


S11B

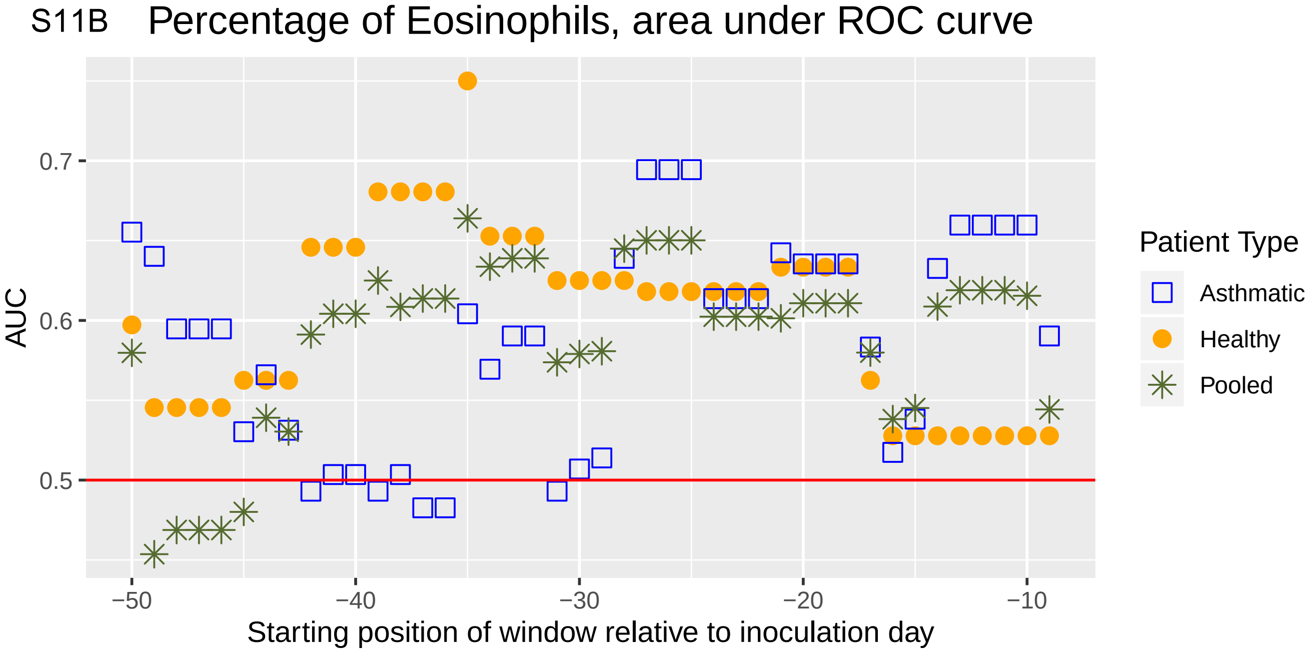

Percentage of Eosinophils, area under ROC curve
0.7
0.6
0.5
AUC
−50
−40
−30
−20
−10
Starting position of window relative to inoculation day
Patient Type
Asthmatic
Healthy
Pooled

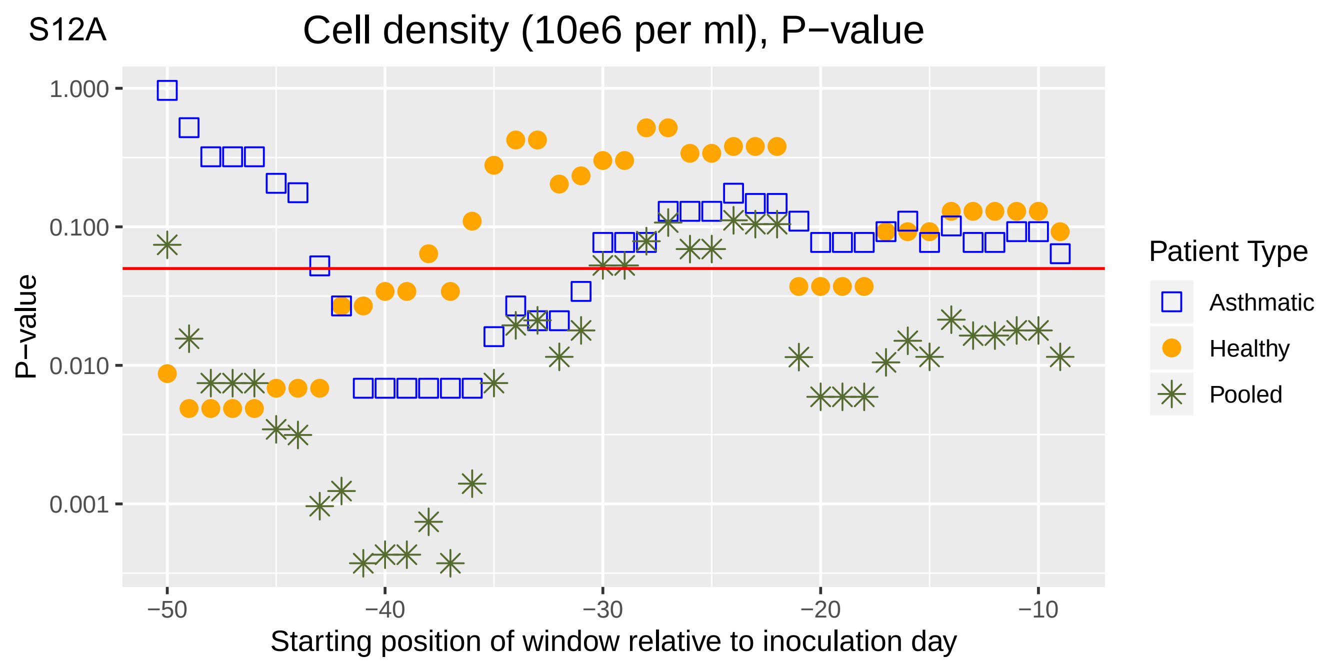
S12A
Cell density (10e6 per ml), P−value
P−value
1.000
0.100
0.010
0.001
−50
−40
−30
−20
−10
Starting position of window relative to inoculation day
Patient Type
Asthmatic
Healthy
Pooled

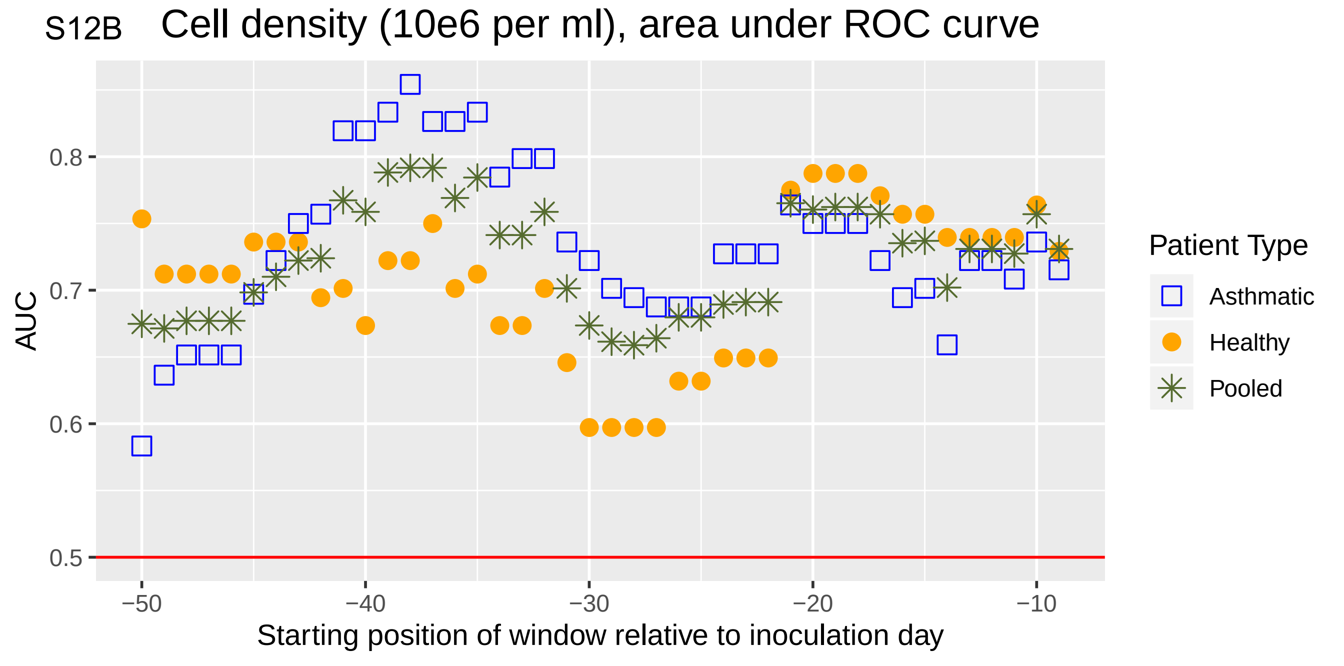
S12B
Cell density (10e6 per ml), area under ROC curve
AUC
0.8
0.7
0.6
0.5
−50
−40
−30
−20
−10
Starting position of window relative to inoculation day
Patient Type
Asthmatic
Healthy
Pooled

S13A

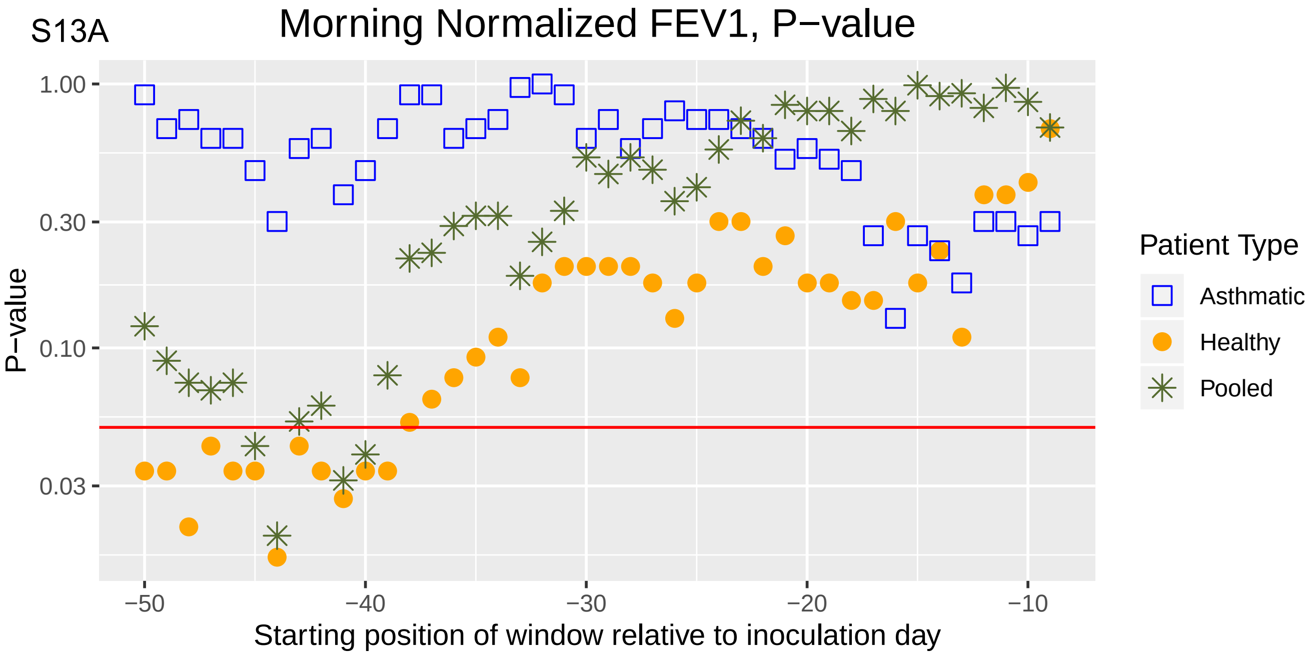

Morning Normalized FEV1, P−value
P−value
1.00
0.30
0.10
0.03
−50
−40
−30
−20
−10
Starting position of window relative to inoculation day
Patient Type
Asthmatic
Healthy
Pooled


S13B

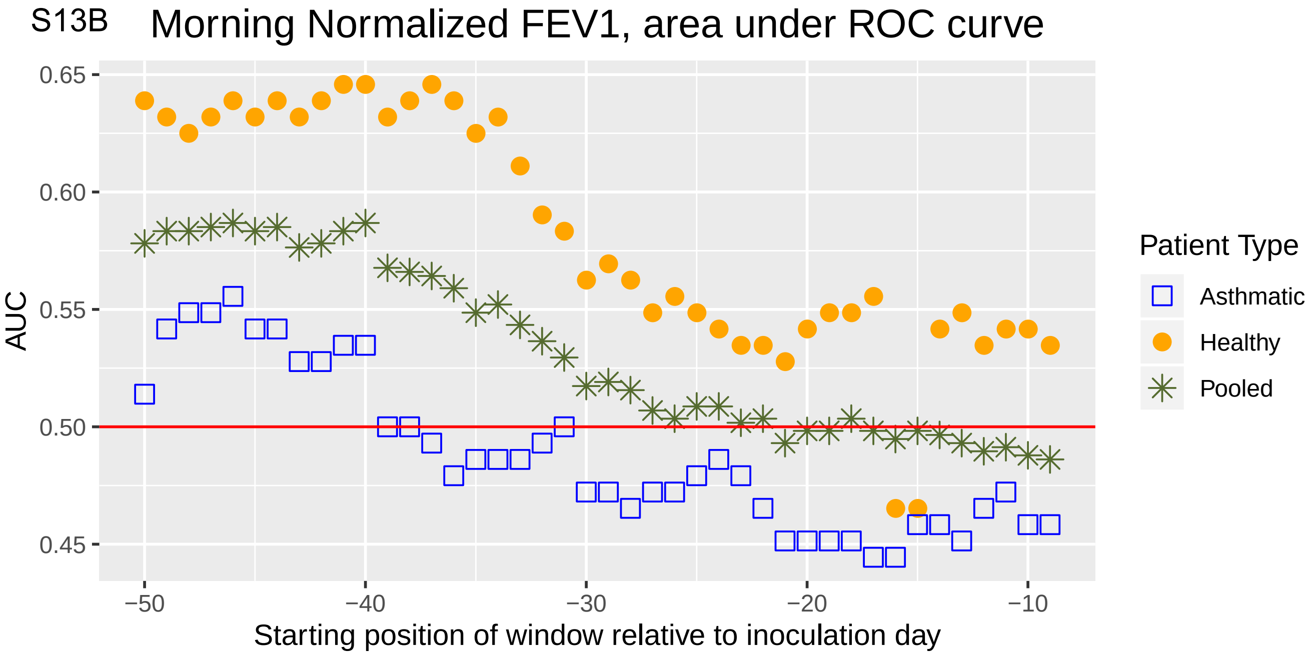

Morning Normalized FEV1, area under ROC curve
AUC
0.65
0.60
0.55
0.50
0.45
−50
−40
−30
−20
−10
Starting position of window relative to inoculation day
Patient Type
Asthmatic
Healthy
Pooled

S14A

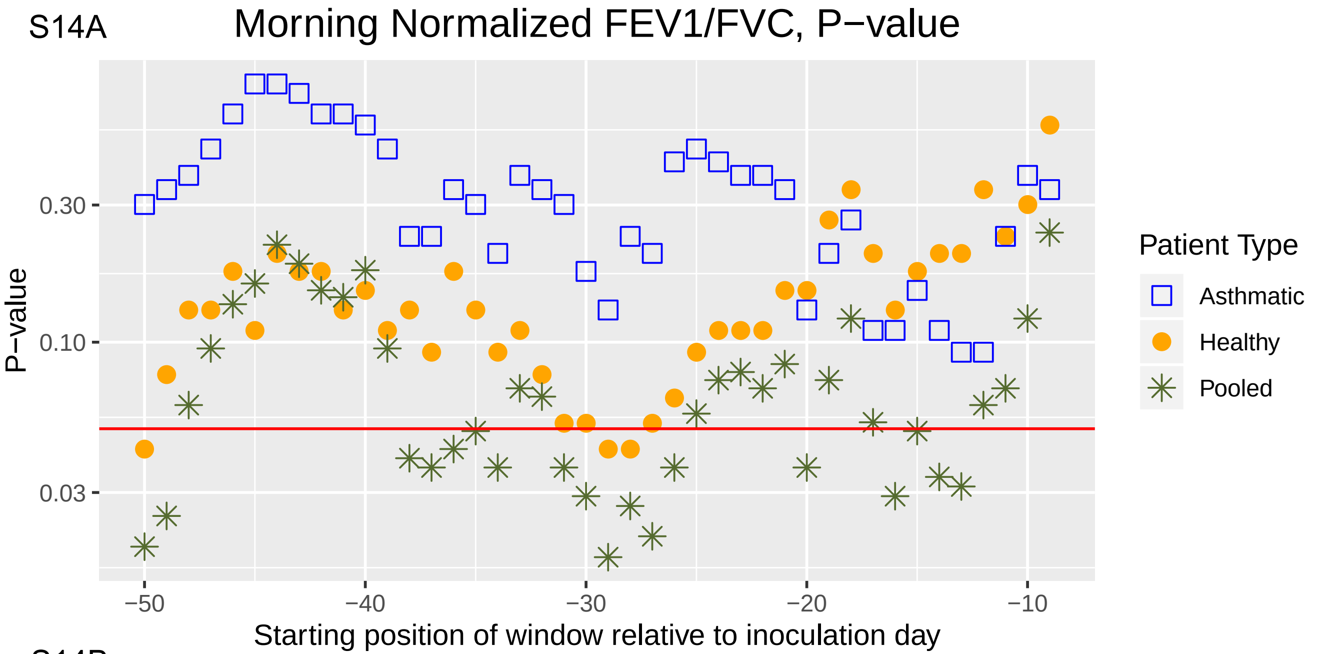

Morning Normalized FEV1/FVC, P−value
P−value
0.30
0.10
0.03
−50
−40
−30
−20
−10
Starting position of window relative to inoculation day
Patient Type
Asthmatic
Healthy
Pooled


S14B

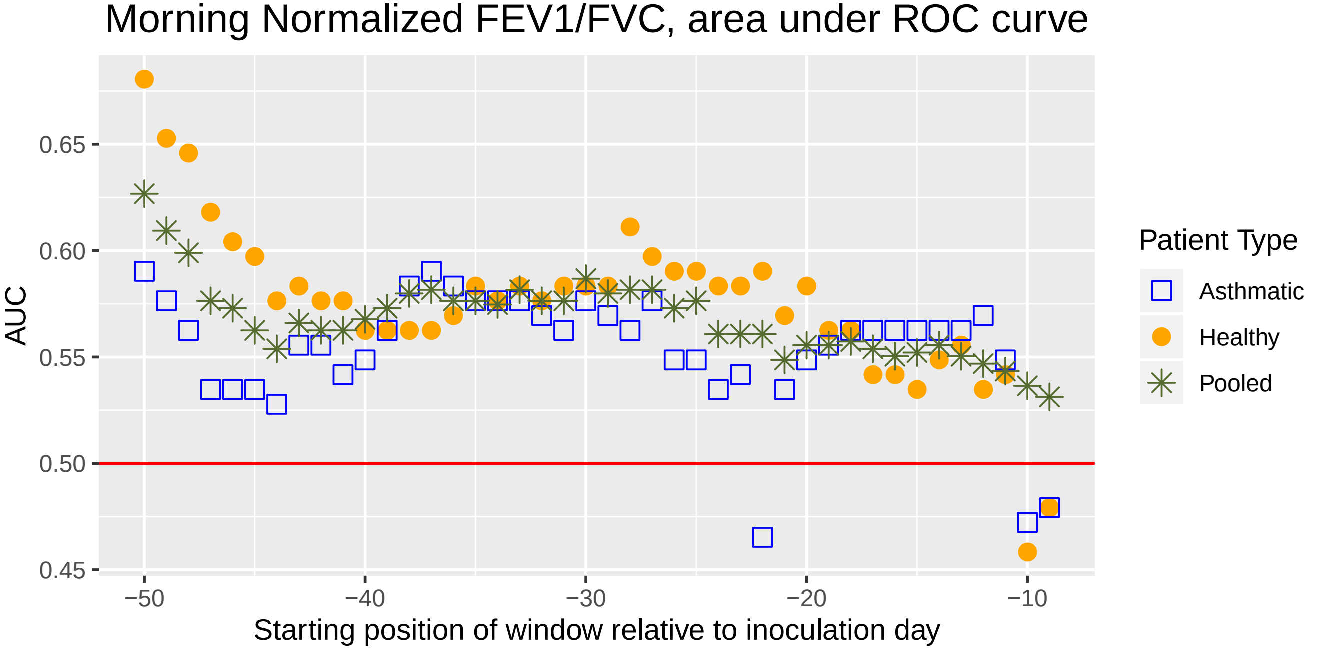

Morning Normalized FEV1/FVC, area under ROC curve
AUC
0.65
0.60
0.55
0.50
0.45
−50
−40
−30
−20
−10
Starting position of window relative to inoculation day
Patient Type
Asthmatic
Healthy
Pooled

S15A

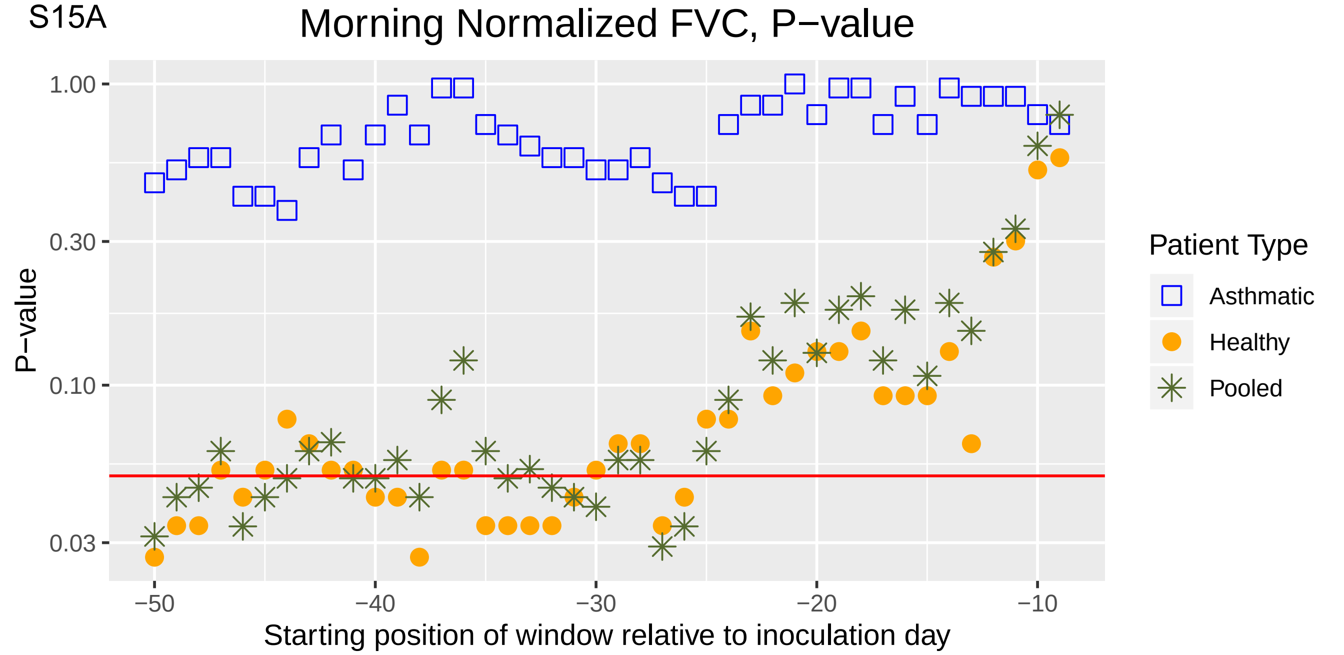
Morning Normalized FVC, P−value
P−value
1.00
0.30
0.10
0.03
−50
−40
−30
−20
−10
Starting position of window relative to inoculation day
Patient Type
Asthmatic
Healthy
Pooled

S15B

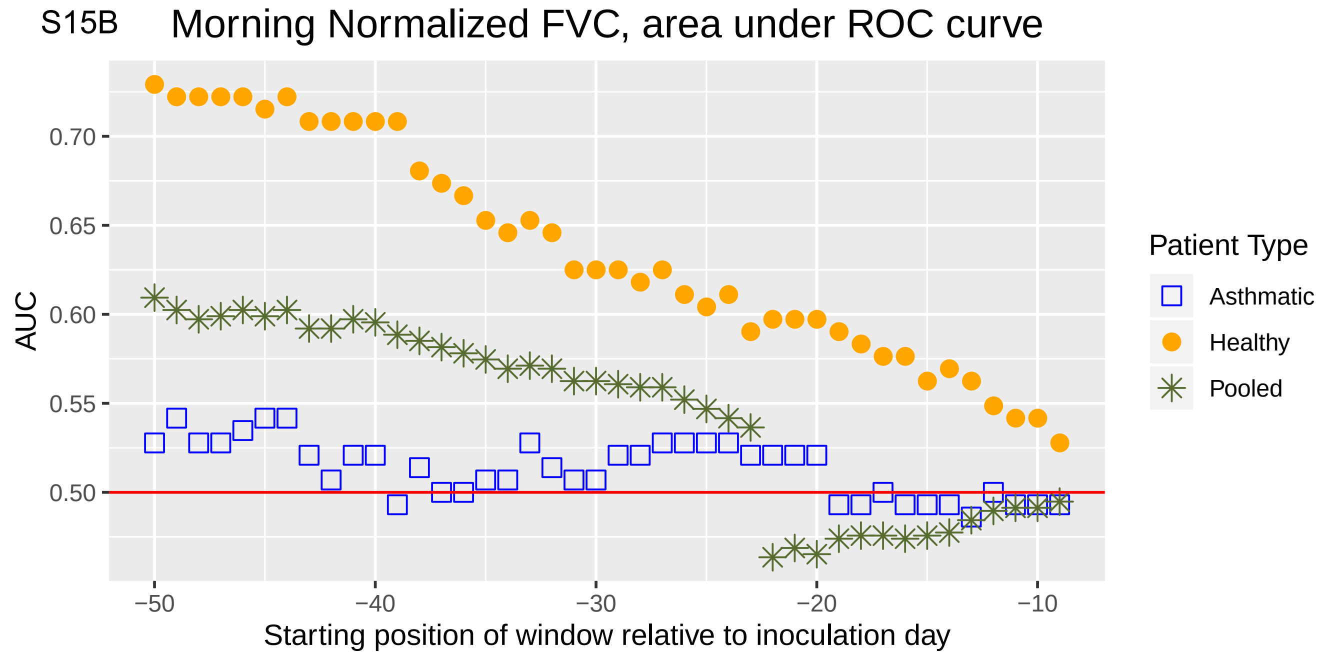
Morning Normalized FVC, area under ROC curve
AUC
0.70
0.65
0.60
0.55
0.50
−50
−40
−30
−20
−10
Starting position of window relative to inoculation day
Patient Type
Asthmatic
Healthy
Pooled

S16A

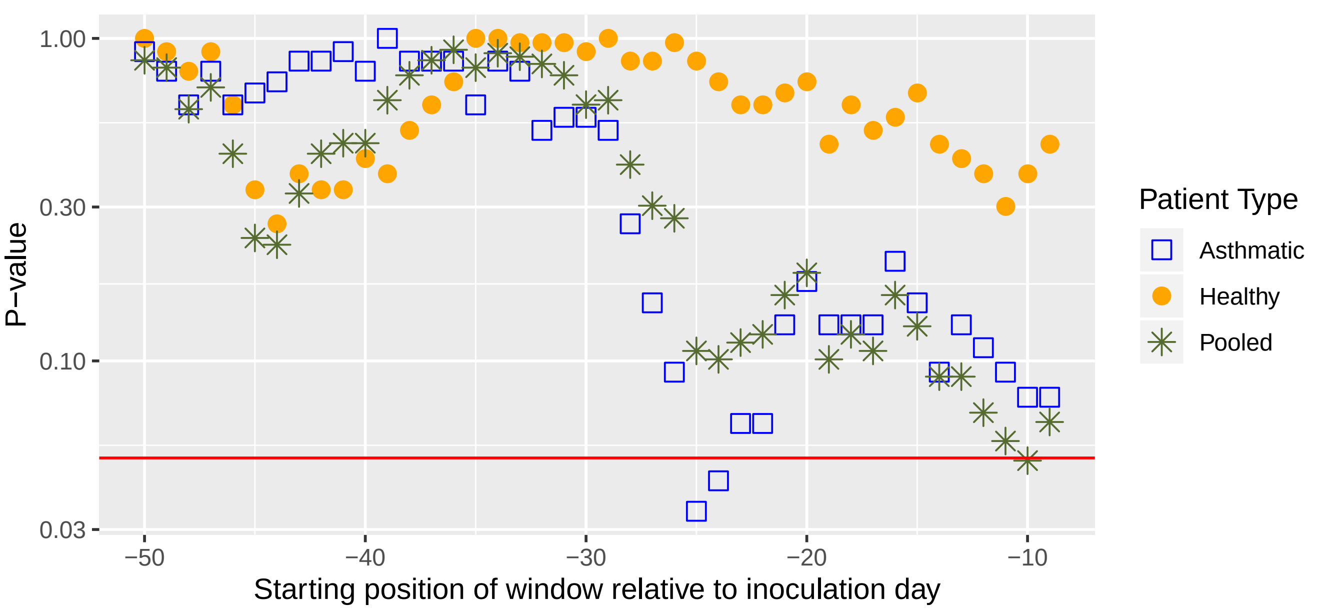

Morning PEF (% pred.), P−value
P−value
1.00
0.30
0.10
0.03
−50
−40
−30
−20
−10
Starting position of window relative to inoculation day
Patient Type
Asthmatic
Healthy
Pooled


S16B

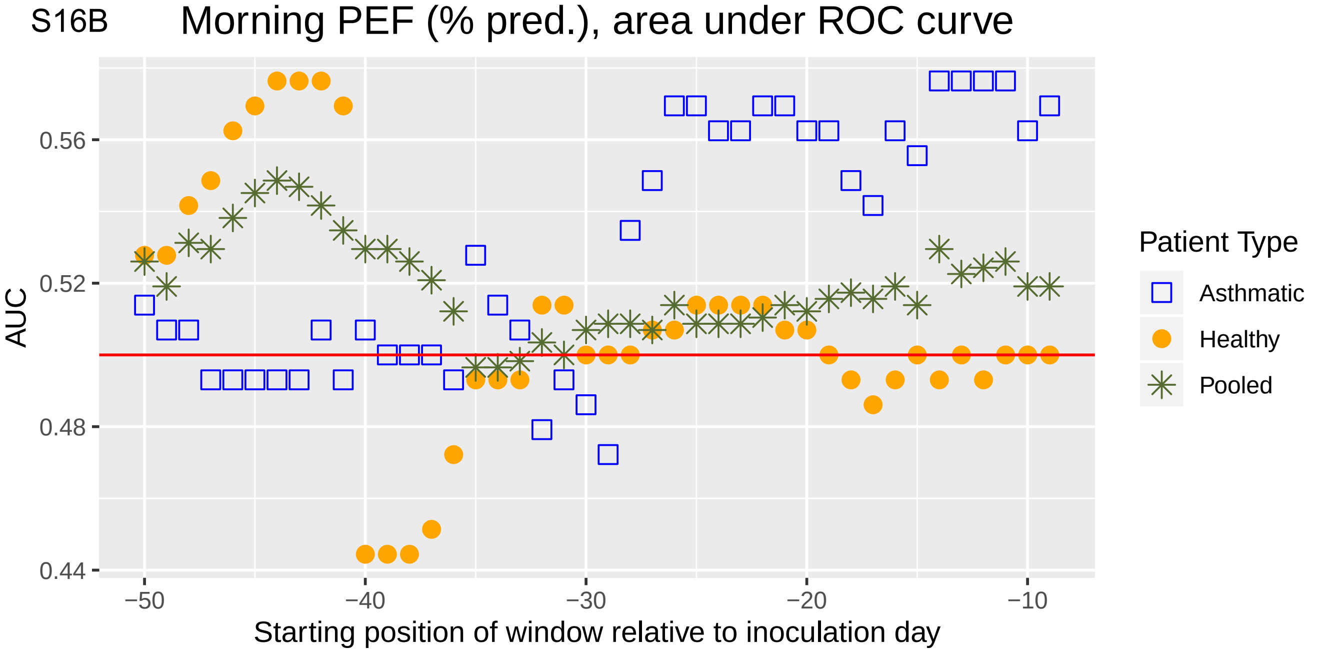

Morning PEF (% pred.), area under ROC curve
AUC
0.56
0.52
0.48
0.44
−50
−40
−30
−20
−10
Starting position of window relative to inoculation day
Patient Type
Asthmatic
Healthy
Pooled

S17A

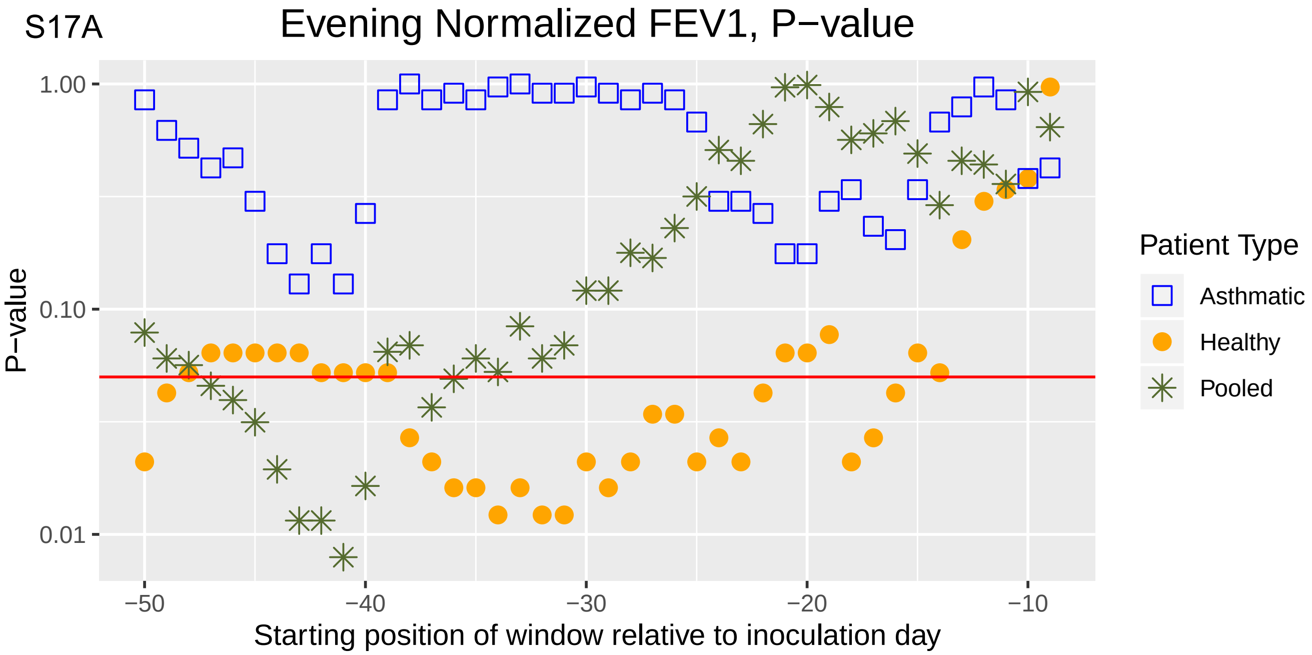
Evening Normalized FEV1, P−value
P−value
1.00
0.10
0.01
−50
−40
−30
−20
−10
Starting position of window relative to inoculation day
Patient Type
Asthmatic
Healthy
Pooled

S17B

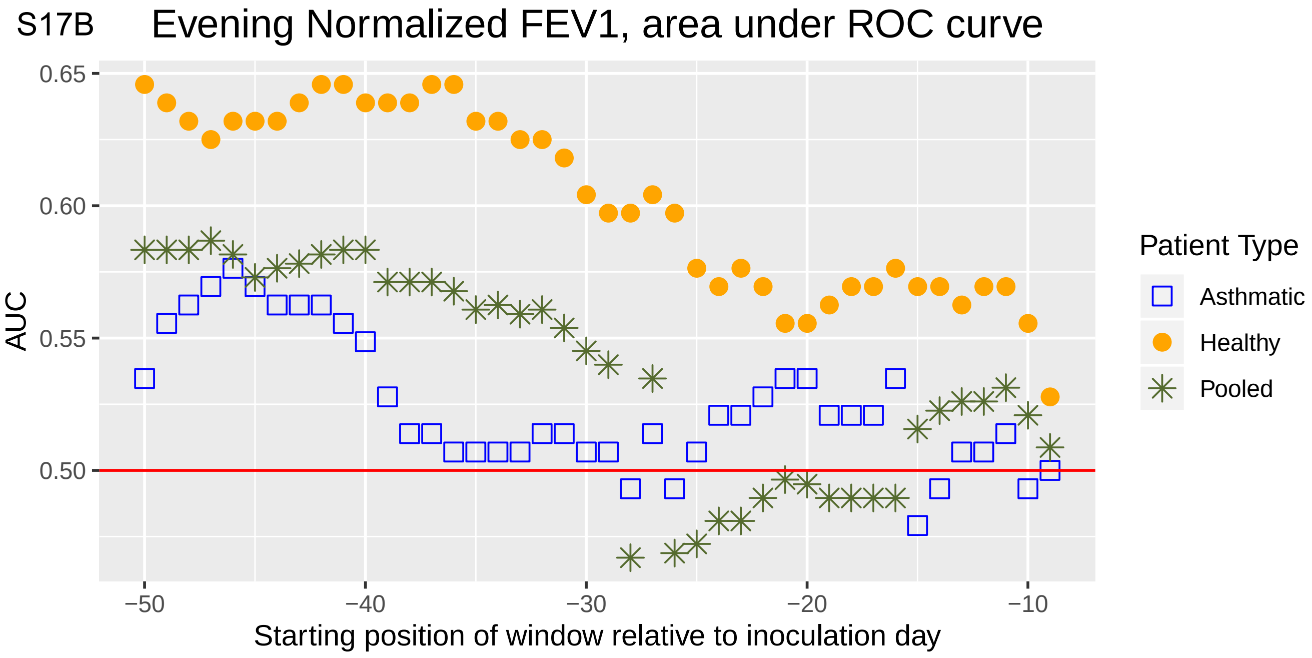
Evening Normalized FEV1, area under ROC curve
AUC
0.65
0.60
0.55
0.50
−50
−40
−30
−20
−10
Starting position of window relative to inoculation day
Patient Type
Asthmatic
Healthy
Pooled

S18A

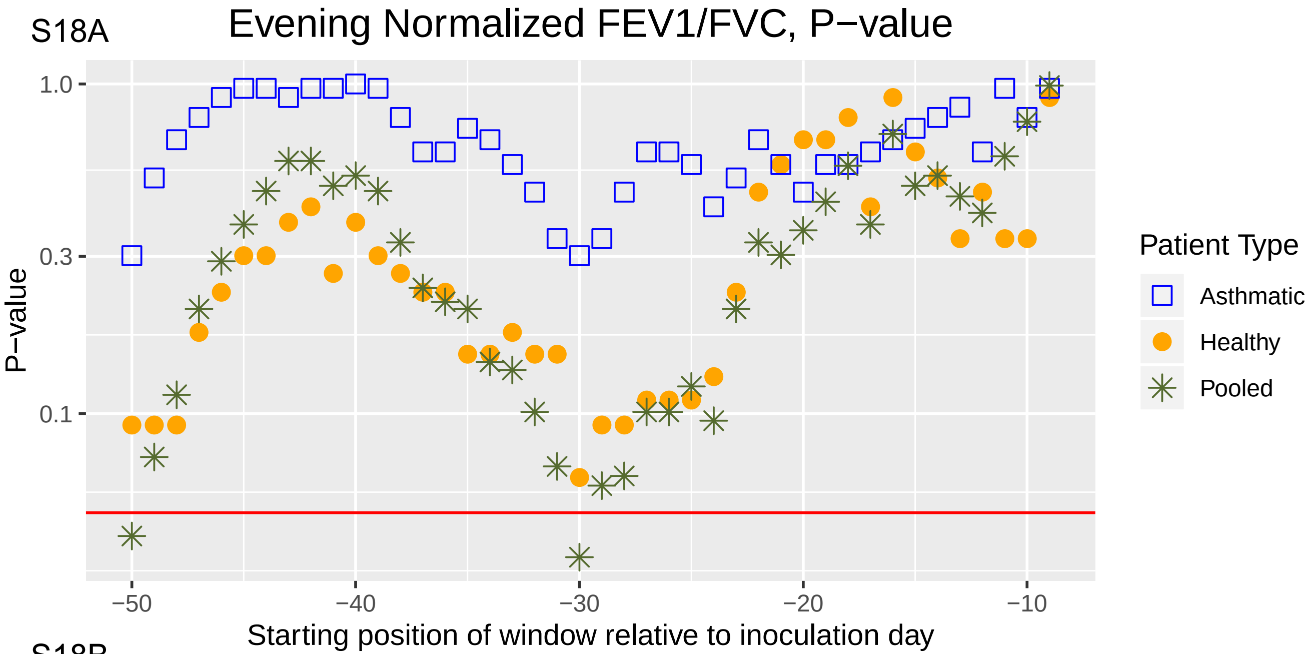
Evening Normalized FEV1/FVC, P−value
P−value
1.0
0.3
0.1
−50
−40
−30
−20
−10
Starting position of window relative to inoculation day
Patient Type
Asthmatic
Healthy
Pooled

S18B

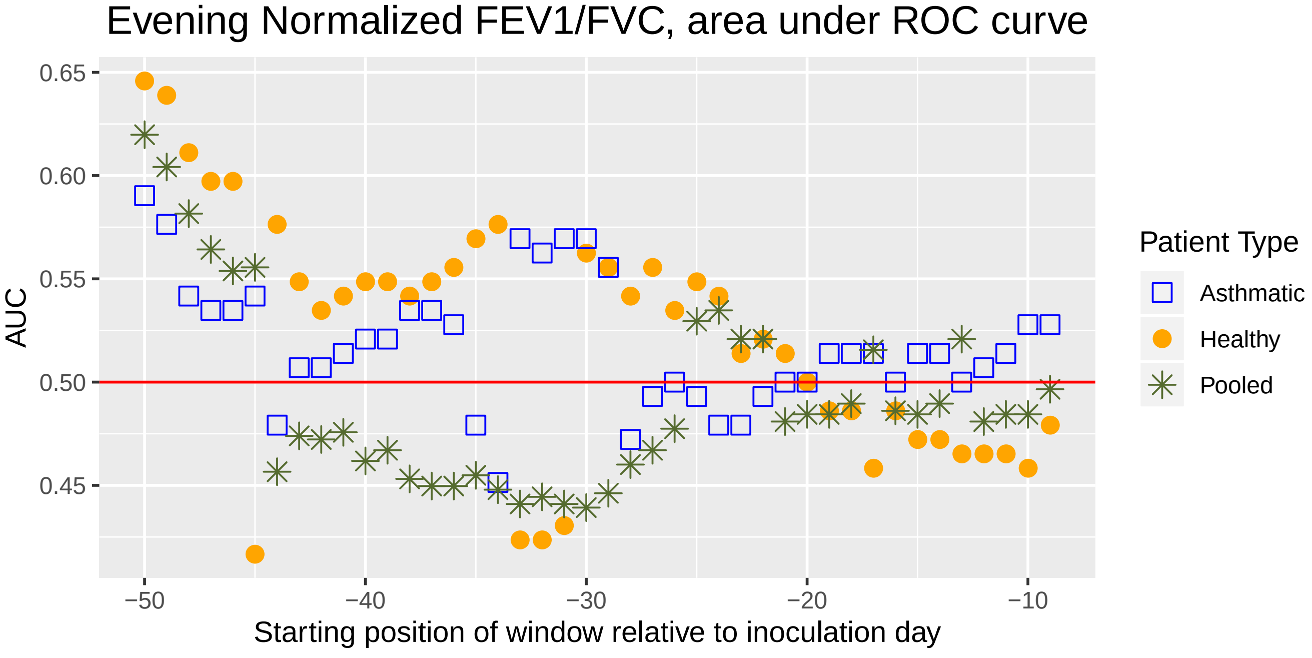
Evening Normalized FEV1/FVC, area under ROC curve
AUC
0.65
0.60
0.55
0.50
0.45
−50
−40
−30
−20
−10
Starting position of window relative to inoculation day
Patient Type
Asthmatic
Healthy
Pooled

S19A

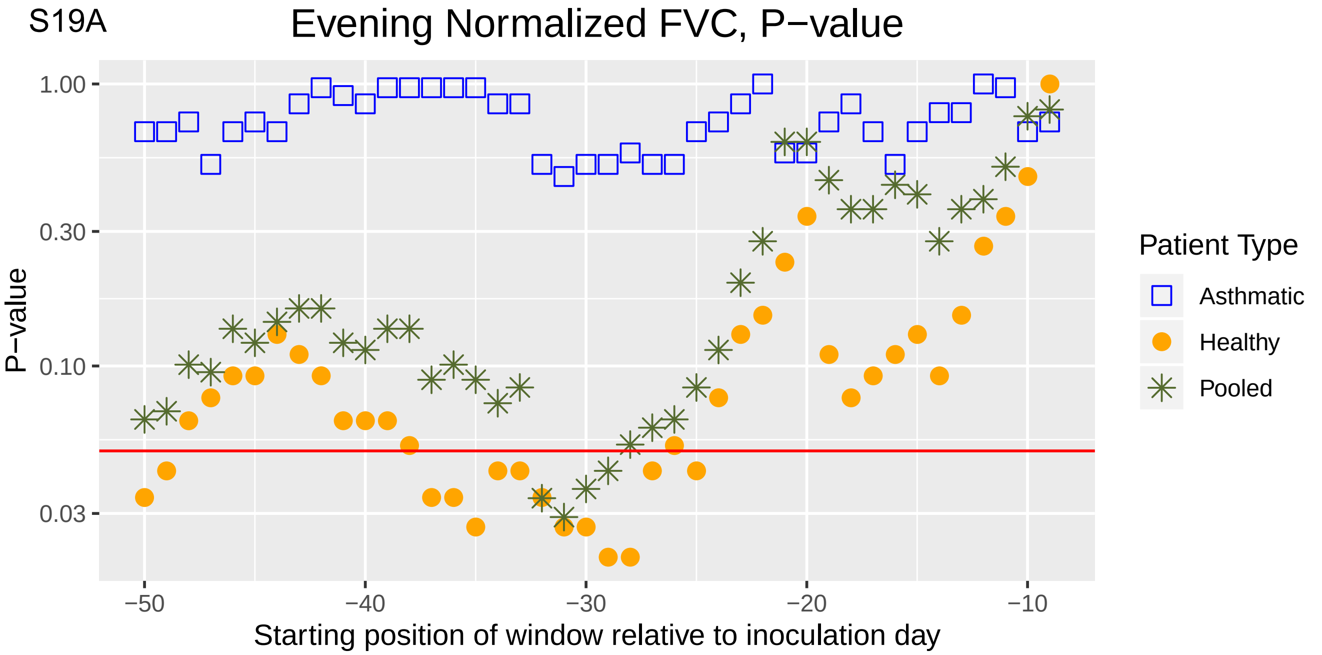

Evening Normalized FVC, P−value
P−value
1.00
0.30
0.10
0.03
−50
−40
−30
−20
−10
Starting position of window relative to inoculation day
Patient Type
Asthmatic
Healthy
Pooled


S19B

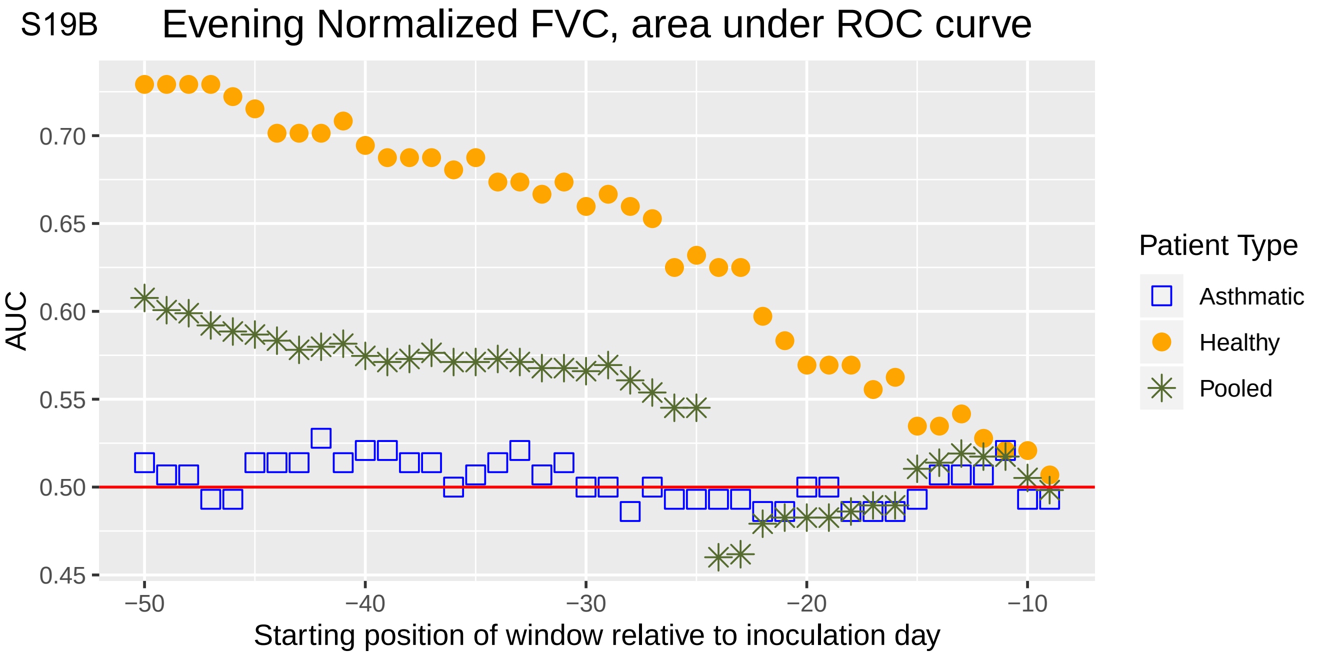

Evening Normalized FVC, area under ROC curve
AUC
0.70
0.65
0.60
0.55
0.50
0.45
−50
−40
−30
−20
−10
Starting position of window relative to inoculation day
Patient Type
Asthmatic
Healthy
Pooled

S20A

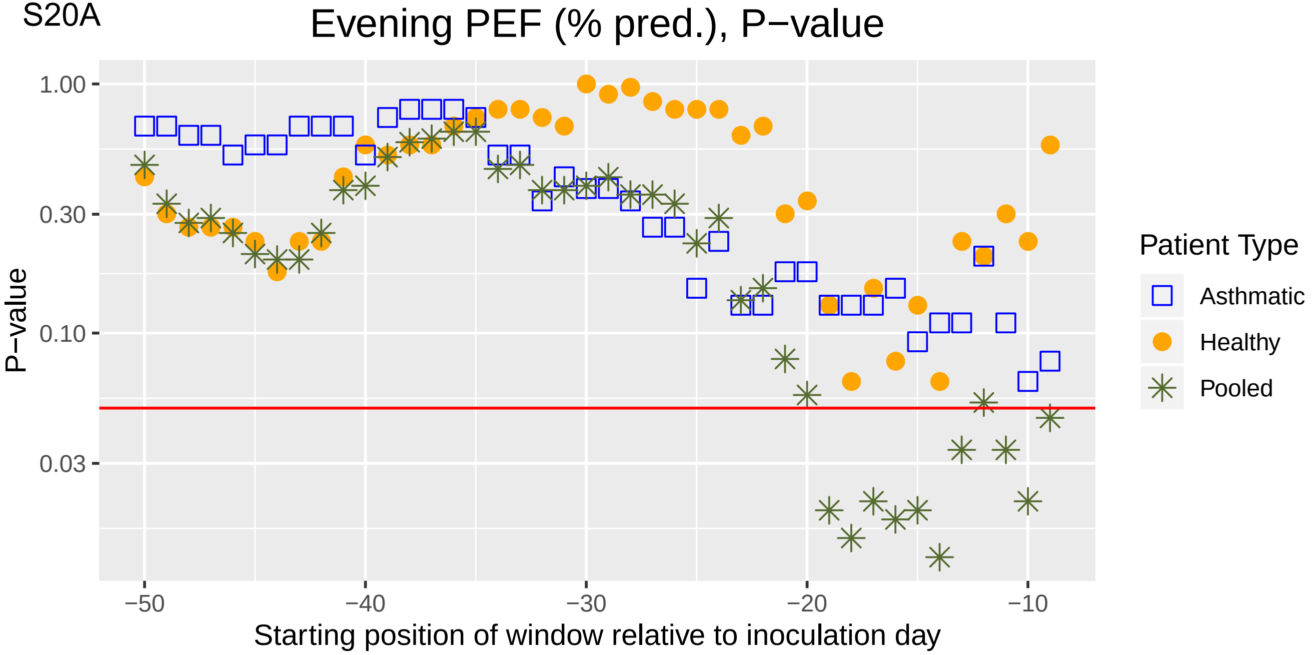
Evening PEF (% pred.), P−value
P−value
1.00
0.30
0.10
0.03
−50
−40
−30
−20
−10
Starting position of window relative to inoculation day
Patient Type
Asthmatic
Healthy
Pooled

S20B

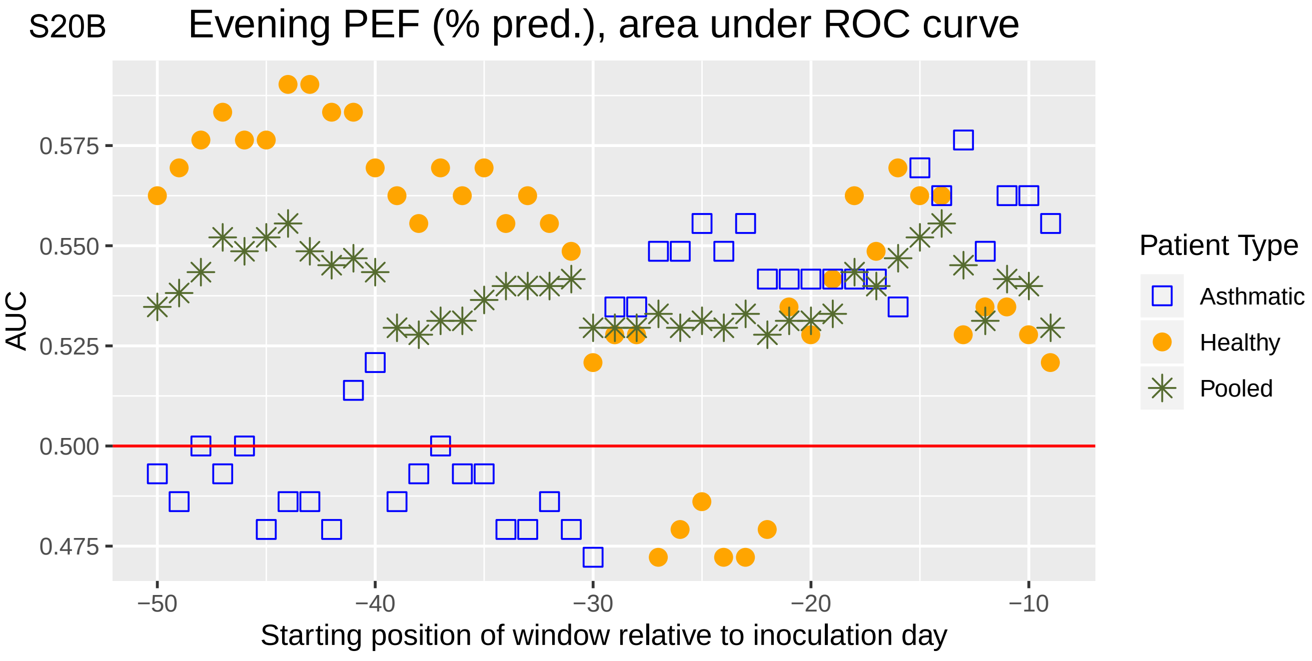
Evening PEF (% pred.), area under ROC curve
AUC
0.575
0.550
0.525
0.500
0.475
−50
−40
−30
−20
−10
Starting position of window relative to inoculation day
Patient Type
Asthmatic
Healthy
Pooled
